# Comparison of COVID-19 and Influenza-Related Outcomes in the United States during Fall-Winter 2022-2023

**DOI:** 10.1101/2023.09.08.23295262

**Authors:** Hagit Kopel, Alina Bogdanov, Jessamine Winer-Jones, Christopher Adams, Isabelle Winer, Mac Bonafede, Van Hung Nguyen, James A. Mansi

## Abstract

**Background:** Three years into the pandemic, SARS-COV-2 remains a significant burden in comparison to other respiratory illnesses; however, many of the monitoring tools available during the early phase of the COVID-19 pandemic have been phased out, making it more difficult to track the current burden of outpatient medical encounters and hospitalizations, especially for at-risk groups. The objective of this analysis was to characterize the frequency and severity of medically-attended COVID-19 and influenza during peak influenza activity in the pediatric (0-17), adult (18-64), and older adult (65+) populations and characterize the prevalence of underlying medical conditions among patients hospitalized with COVID-19.

**Methods:** This was a cross-sectional analysis of individuals in the Veradigm Health Insights EHR Database linked to Komodo claims data with a medical encounter of claim between October 1, 2022, and March 31, 2023. We captured age, sex, and underlying medical conditions associated with higher risk for severe COVID-19 during a 12-month baseline period. We identified patients with medical encounters with a diagnosis of COVID-19 or influenza between October 1, 2022, and March 31, 2023, and stratified them into 5 mutually exclusive categories based on the highest level of care received with that diagnosis during the season (intensive care unit [ICU] > hospitalization without ICU > emergency department > urgent care > other outpatient).

**Results:** Among the 23,526,196 individuals in the dataset, 5.0% had a COVID-19-related medical encounter, and 3.0% had an influenza-related medical encounter during the 6 month observation period. The incidence of hospitalizations with a COVID-19 diagnosis was 4.6 times higher than the incidence of hospitalizations with an influenza diagnosis. Hospitalizations with COVID-19 were higher in all age groups. Nearly all adults hospitalized with COVID-19 had at least one underlying medical condition, but 25.8% of 0-5-year-olds and 18.3% of 6-17-year-olds had no underlying medical conditions.

**Conclusions:** COVID-19 continues to place a heavy burden on the United States healthcare system and was associated with more medical encounters in all age groups, including hospitalizations, than influenza during a 6-month period that included the 2022-2023 peak influenza activity.

## Introduction

Severe acute respiratory syndrome coronavirus 2 (SARS-CoV-2), the virus responsible for the coronavirus 2019 (COVID-19) respiratory infection, was first detected in Wuhan, China, in December of 2019.^1^ It quickly spread globally and has proven to be a persistent public health threat due in part to variants with enhanced transmissibility and pathogenicity.^2^ In addition, previously widespread variants are rapidly displaced by new variants and sub-variants that more effectively evade existing natural and vaccine immunity.^3^ As a result, despite widespread immune exposure to the SARS-COV-2 virus, test positivity for COVID-19 remains high and exceeded test positivity for influenza and respiratory syncytial virus during the majority of the last 12 months in the US.^4^ Infection may result from a short but intense exposure or following prolonged or repeated exposure to a smaller dose over time, such as when SARS-CoV-2 is introduced within a household. One study found that while 75% of children had asymptomatic infection, the secondary attack rate in the home was 57.7%.^5^

Although COVID-19 is no longer considered a public health emergency,^6^ it continues to be the leading respiratory infectious disease causing hospitalizations in the United States.^4,7,8^ Previous research has shown that COVID-19 disproportionately affects older adults and those with underlying medical conditions, placing them at higher risk for COVID-19-related morbidity and mortality.^9–11^ In addition, infection with SARS-CoV-2 is associated with post-acute sequelae (i.e., long COVID) that include a wide range of health issues that can occur even among patients who experienced a mild acute infection and can persist for years following initial infection.^12–14^ According to the Household Pulse Survey, 27.6% of adults who had COVID report experiencing long COVID.^15^ Studies have shown that patients hospitalized with COVID-19 were over twice as likely to develop hypertension compared with patients who had influenza,^16^ and long Covid was associated with a higher burden of disability than either heart disease or cancer.^13^

Vaccination has been shown to reduce COVID-19 incidence and severity and is associated with a 30-40% lower odds of long COVID;^17,18^ however, vaccine efficacy wanes with viral mutation and time since most recent dose.^19,20^ Repeat doses and new vaccine formulations can help restore waning immunity and provide protection against the newer variants.^21,22^ Yet vaccination rates have decreased with each successive dose.^23^ During the 2022-2023 season, vaccines were developed to counter waning immunity and broaden protection against emerging variants.

Unfortunately, vaccination rates were low, with only ∼17% of those eligible having received bivalent vaccination as of August 2023,^24^ which was substantially lower than the annual influenza vaccination during the same period. ^25^ This raises public health concerns about suboptimal protection against COVID-19, particularly during the most recent influenza season.

The objective of this analysis was to characterize the frequency and severity of medically-attended COVID-19 and influenza during a 6-month period (October 1, 2022 through March 31, 2023) that included peak influenza activity in the pediatric (0-17), adult (18-64), and older adult (65+) populations. A secondary objective was to characterize the prevalence of underlying medical conditions among patients hospitalized with COVID-19.

## Methods

### Study Design and Data Sources

We conducted a cross-sectional analysis of COVID-19 and influenza medical encounters from October 1, 2022, through March 31, 2023, a six-month period roughly covering the peak of the 2022-2023 influenza season. This study leveraged electronic health records (EHRs) from the Veradigm Health Insights Database linked to administrative claims data from Komodo Health’s Healthcare Map. The EHR dataset consists of patient records sourced from ambulatory/outpatient primary care and specialty settings. The insurance claims data contains inpatient, outpatient, and pharmacy claims, and only closed claims were used for this study. Patient-level files in each data source are linked by de-identified tokens created by Datavant to create a final de-identified dataset that contains no protected health information. The linked dataset has been determined to be statistically de-identified through a formal determination by a qualified expert as defined in Section §164.514(b)(1) of the Health Insurance Portability and Accountability Act Privacy Rule.

As a noninterventional, retrospective database study using a certified Health Insurance Portability and Accountability Act-compliant deidentified research database, approval by an institutional review board was not required.

### Cohort Construction

This study included all individuals in the linked dataset who had continuous claims enrollment with both pharmacy and medical benefits between October 1, 2022, and March 31, 2023 (study period), and at least one record of EHR or claims activity during that period. Codes used to identify COVID-19 and influenza cases are reported in Supplementary File 1.

### Patient Characteristics

For each patient, we recorded age and sex at the start of the study period. Age is reported categorically in the following groups.

We also captured underlying medical conditions identified by the CDC as associated with higher risk for severe COVID-19 that were documented in the patient record during the 12 months prior to October 1, 2022.^26^ The underlying medical conditions of interest included in this analysis were attention deficit and hyperactivity disorder (ADHD), asthma, cancer, cerebral palsy, cerebrovascular disease, chronic kidney disease, chronic liver disease, chronic lung disease, congenital malformation, cystic fibrosis, dementia (only in individuals 18+), diabetes (type 1 and type 2), disability, Down syndrome, heart disease, human immunodeficiency virus (HIV), hypertension, use of immunosuppressive medications (including topical corticosteroids), mental health conditions, musculoskeletal conditions, neurologic conditions, obesity (body mass index > 30), other immunocompromised condition, physical inactivity, pregnancy (only in individuals 18+), smoking (current and former; only in individuals 18+), solid organ transplant, stem cell transplant, and tuberculosis. Code sets are listed in Supplementary File 1.

### Study Outcomes

For each patient, we looked for encounters with a diagnosis of COVID-19 during the study period and assigned patients to one of 6 mutually exclusive categories: hospitalized with intensive care unit (ICU) admission, hospitalization without ICU, emergency department (ED), urgent care, outpatient visit (includes but not limited to primary care visits, office visits, and telemedicine visits), or none. A patient could be included in both the COVID-19 outcomes and the influenza outcomes if they had a diagnosis of each during the study period. However, if patients had more than one encounter with a COVID-19 or influenza diagnosis during the study period, they were assigned to a single category based on the following hierarchy: ICU > hospitalization without ICU> ED > urgent care > outpatient. Code sets for COVID-19 and influenza are listed in Supplementary File 1.

This analysis was repeated for influenza diagnoses during the study period. While patients could be assigned to only one COVID-19 category and only one influenza category, they could be assigned to both a COVID-19 category and an influenza category if they had a diagnosis of each during the study period.

### Data Analysis

All results are reported descriptively. Incident rates were calculated per 100,000 individuals over the 6 month study period during which patients had continuous claims enrollment. All variables are categorial and reported as counts and percentages. The data file was constructed and analyzed using SAS V9.4 (SAS, Cary, NC).

## Results

The final dataset included 23,526,196 individuals with an insurance claim or medical record between October 1, 2022, and March 31, 2023. Overall, 58.0% of the included population were female, and 65.7% were 18 to 64 years old at the start of the season (Table 1). In the 12 months preceding the study period, 35.5% of the population had a record of use of immunosuppressive medications (including prednisone and topical corticosteroids), 29.0% had a diagnosis of hypertension, and 21.0% were obese.

**Table 1.**
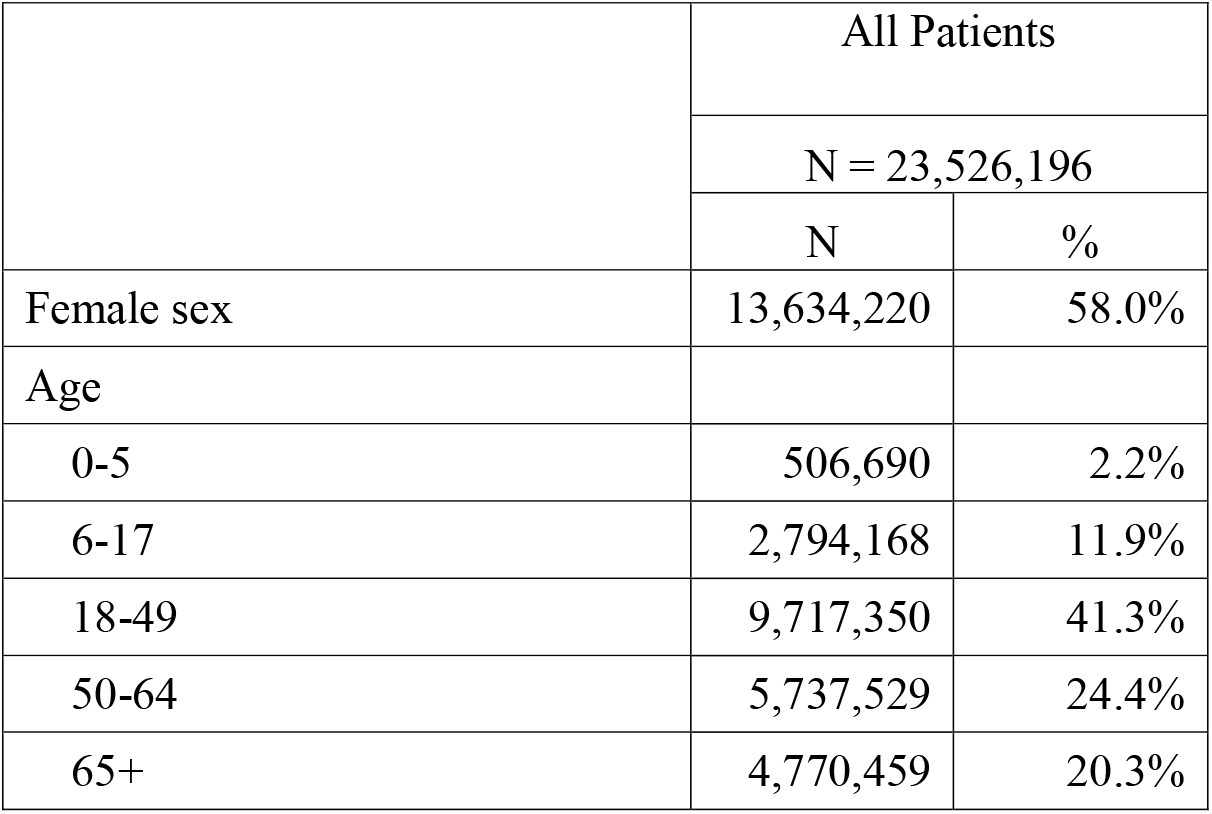
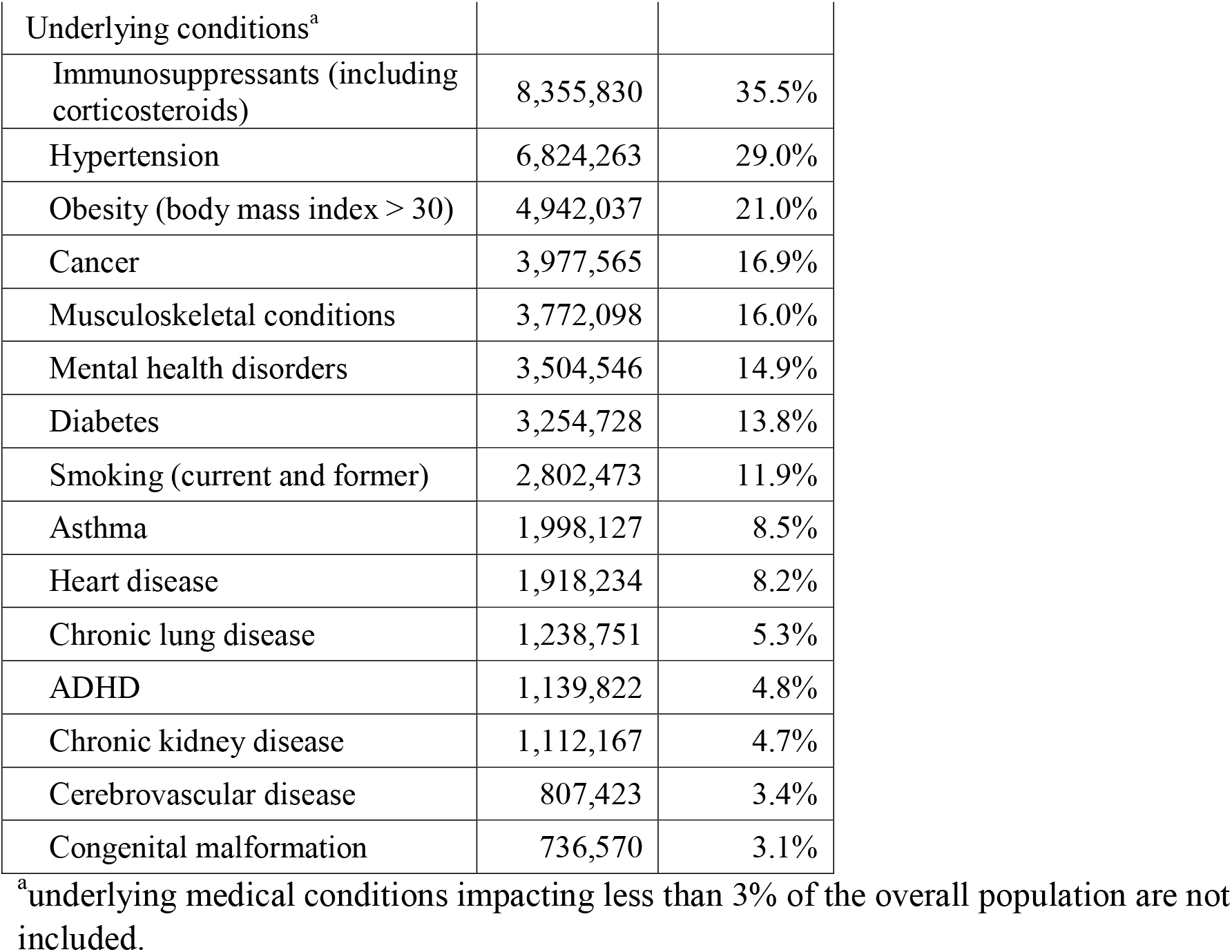
Population Characteristics.

### Incidence Rate of COVID-19 or Influenza Related Outcomes

During the study period, 5.0% (N = 1,179,960) of the study population had a medical encounter with a COVID-19 diagnosis, and 3.0% (N = 698,002) had a medical encounter with an influenza diagnosis (Table 2). The majority of both COVID-19 and influenza encounters occurred in the urgent care setting (53.5% and 69.7%, respectively). However, COVID-19 was associated with more severe outcomes as 7.0% of COVID-19 visits occurred in the hospital non-ICU setting compared to 2.6% of influenza visits, and 1.0% of COVID-19 visits occurred in the ICU setting compared to 0.4% of influenza visits.

**Table 2.** COVID-19 and influenza medical encounters between October 1, 2022, and March 31, 2023.

|  | COVID-19 |  |  | Influenza |  |  | Incidence Ratio of COVID-19 to Influenza |
| --- | --- | --- | --- | --- | --- | --- | --- |
|  | N = 1,179,960 |  |  | N = 698,002 |  |  |  |
|  | N | %<sup>a</sup> | Incidence<sup>b</sup> | N | %<sup>a</sup> | Incidence<sup>b</sup> |  |
| Outpatient visits (other) | 283,213 | 24.0% | 1,204 | 50,835 | 7.3% | 216 | 5.6 |
| Urgent care visit | 631,356 | 53.5% | 2,684 | 486,406 | 69.7% | 2,068 | 1.3 |
| ED visit | 171,503 | 14.5% | 729 | 140,200 | 20.1% | 596 | 1.2 |
| Hospitalization<sup>c</sup> | 82,234 | 7.0% | 350 | 18,032 | 2.6% | 77 | 4.6 |
| ICU | 11,654 | 1.0% | 50 | 2,529 | 0.4% | 11 | 4.6 |

ICU, intensive care unit; ED, Emergency department
<sup>a</sup>Percents calculated per total number of COVID-19 or influenza cases
<sup>b</sup>Incidence calculated as per 100,000 people per 6-month season among the 23,526,196 individuals with continuous claims enrollment for that period,
<sup>c</sup>Not including ICU

Incidence rates of all outcomes were higher among those with COVID-19 than among those with influenza (Table 2). For example, the incidence of hospitalization with or without ICU was 4.6 times higher for COVID-19 than for influenza.

### COVID-19 and Influenza-Related Hospitalizations by Age

Overall, between October 1, 2022, and March 31, 2023, we identified 93,888 individuals who were hospitalized with COVID-19 (with or without ICU admission) and 20,561 individuals who were hospitalized with influenza (Table 2). Roughly 12% of hospitalizations with either COVID-19 or influenza included ICU admission. In all age groups, a greater number of patients were hospitalized with COVID-19 than with influenza during the study period (Table 3). Notably, there were 5.6 times as many patients aged 18-49 and 4.2 times as many patients aged 50-64 hospitalized with COVID-19 than hospitalized with influenza.

**Table 3.** Hospitalizations* with a diagnosis of COVID-19 or influenza across age groups between October 1, 2022, and March 31, 2023.

|  | COVID-19<br>N = 93,888<br>N | Influenza<br>N = 20,561<br>N | Count Ratio<br>of COVID-19 vs<br>Influenza |
| --- | --- | --- | --- |
| **Pediatrics** |  |  |  |
| 0-5 | 706 | 564 | 1.3 |
| 6-17 | 1,529 | 1,260 | 1.2 |
| **Adults** |  |  |  |
| 18-49 | 26,242 | 4,693 | 5.6 |
| 50-64 | 22,947 | 5,529 | 4.2 |
| **Older Adults** |  |  |  |
| 65+ | 42,464 | 8,515 | 5.0 |

\* Inclusive of intensive care unit admission

### COVID-19 Hospitalizations by Underlying Medical Conditions

Across all age groups, patients with underlying medical conditions were disproportionately represented among patients hospitalized with COVID-19 compared to their prevalence in the dataset. Underlying medical conditions were more common among patients hospitalized with COVID-19 than among the overall population in the dataset (Figure 1 and Supplementary File 2). For example, 64.8% of adults over 65 in the dataset had a prior diagnosis of hypertension, but 85.4% of adults over 65 who were hospitalized with COVID-19 had a prior diagnosis of hypertension. Similarly, 24.9% of adults over 65 in the dataset had a prior diagnosis of heart disease, but 54.0% of adults over 65 who were hospitalized with COVID-19 had a prior diagnosis of heart disease. While the most common underlying medical conditions differed across age groups, the trend of an increased incidence of underlying medical conditions among patients hospitalized with COVID-19 was consistent in all groups. Notably, while in the adult population, 94.4% of patients 18-64 years old and 99.6% of patients 65+ years old hospitalized with COVID-19 had at least one underlying medical condition, in the pediatric population, 25.8% of hospitalized patients 0-5 years old and 18.3% of hospitalized patients 6-17 years old did not have underlying medical conditions.

**Figure 1.**
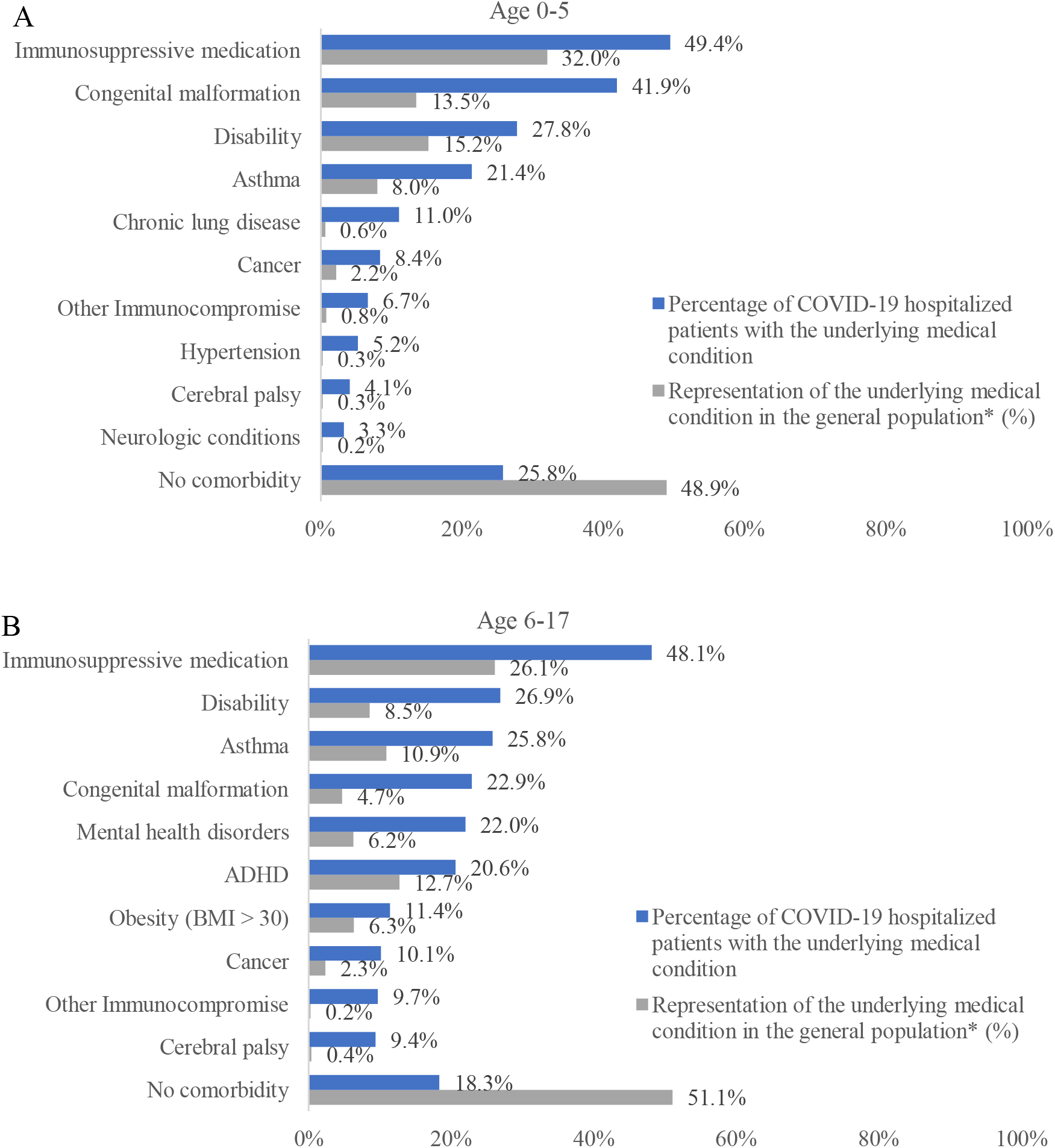
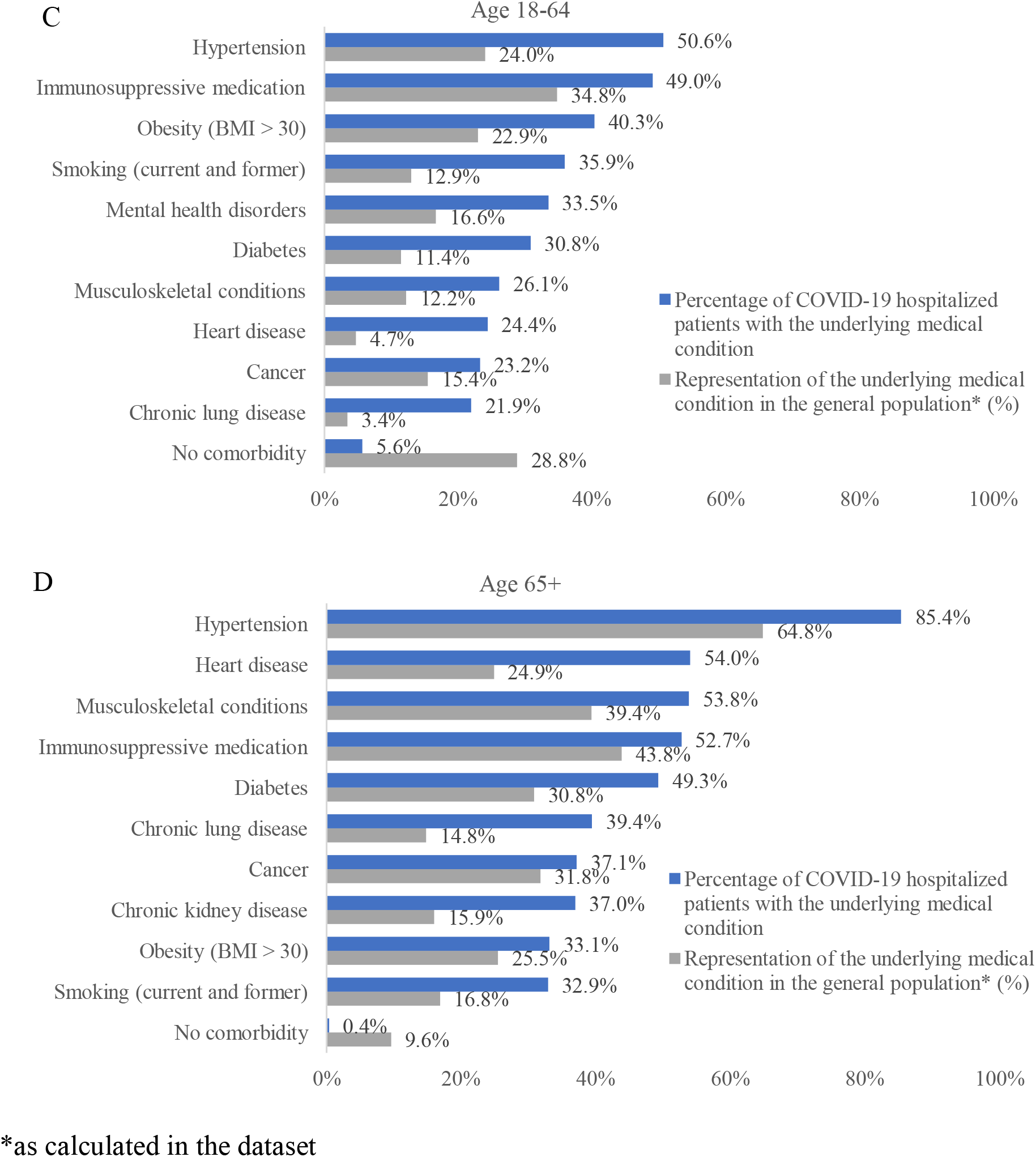
Hospitalizations by underlying medical condition by age (A) 0-5 years old, (B) 6-17 years old, (C) 18-64 years old, and (D) 65+ years old.

## Discussion

In this analysis of over 23 million individuals who used healthcare services between October 1, 2022, through March 31, 2023 (a period that included the peak influenza activity), there were 1.7 times as many patients with a COVID-19-related medical encounter than patients with an influenza-related medical encounter. Hospitalizations, in particular, were more common among patients with COVID-19 than among patients with influenza. Hospitalizations with COVID-19 were more common than hospitalizations with influenza in all age groups, and this difference was particularly striking among adults 18-64 and 65+ years old. Nearly all adults hospitalized with COVID-19 had at least one underlying medical condition associated with increased risk for severe outcomes. Some of the conditions that were most strongly associated with severe COVID-19-related hospitalizations, such as hypertension, diabetes, and obesity, are highly prevalent in the US adult population, including young adults as a 2020 analysis of NHANES found that 75% of all adults including 59% young adults (18-29 years old) had at least one risk factor for severe COVID-19.^27^ Importantly, 26% of those hospitalized between 0-5 years old and 18% of those hospitalized between 6-17 years old had no underlying medical conditions that place them at higher risk of severe COVID-19, making it challenging to predict which children will have severe outcomes from COVID-19.

The trends observed in our analysis are generally consistent with the data available for October 8, 2022, through April 1, 2023, from the CDC’s COVID-NET and FluSurv-NET databases, which track laboratory-confirmed hospitalizations from contributing hospital systems in 13 states.^7,8^ Similar to our study, the incidence of hospitalizations with laboratory-confirmed COVID-19 was markedly higher (∼3 times) than the incidence of hospitalizations with laboratory-confirmed influenza.^7,8^ This difference is smaller than the 4.6 times higher incidence of hospitalization observed in this study but likely reflects the difference in the study methodologies. It is also consistent with the trends observable in the Respiratory Illnesses dashboard developed by Epic research.^4^

In the CDC data for October 8, 2022, through April 1, 2023, roughly 14.6% of hospitalizations with COVID-19 included ICU admission, which is comparable to the 12.4% observed in this study.^7^ In addition, consistent with our analysis, children under 18 hospitalized with COVID-19 at any time were more likely to have no select underlying medical conditions compared to adults over 18 in the CDC data. Previous analysis of the CDC data has found that the annual COVID-19-associated hospitalization rate during 2020-2021 was higher among children <18 years of age than the influenza-associated hospitalization rate during the prior three seasons.^28^ One notable difference between our findings and the CDC data is that in our study, individuals over 65 years old made up only 45.2% of those hospitalized with COVID-19; however, in the CDC data for a similar time period (October 8, 2022, through April 1, 2023) individuals over 65 years old made up only 61.8% of those hospitalized with laboratory-confirmed COVID-19. This difference may be due to differences in the definition of hospitalized with COVID-19, imbalances in who gets tested for COVID-19, and fundamental differences in the sampled population of the two datasets and coverage of the Medicare population. Overall, the CDC data supports our findings that during the most recent influenza season, COVID-19 presented a greater healthcare burden than influenza, regardless of age. Moreover, these data are consistent with other studies comparing the clinical course of COVID-19 and influenza.^29–31^

Like influenza, the incidence of COVID-19 correlates with the arrival of new variants that evade existing immunity;^32,33^ however, the arrival and transmission of new COVID-19 variants does not have an established seasonality. Therefore, it is important to acknowledge that although this study is looking at infections during a period that included the peak influenza activity, COVID-19 diagnoses were still more frequent than influenza diagnoses, particularly among hospitalized patients.

Although vaccination coverage was not measured in this analysis, prior studies have shown that the incidence of any COVID-19 medically-attended outcome is lower among vaccinated individuals compared to unvaccinated individuals.^17^ In addition, timely administration of booster doses has been associated with a lower incidence of COVID-19 during the periods dominated by the Delta and Omicron variants compared to individuals who completed their primary series but did not receive a booster dose.^22^ As new variants emerge, there is a need to assess the effectiveness of recommended vaccines and update them as needed to ensure optimal protection against the circulating variants. In addition, clear evidence-based recommendations and consistent messaging on vaccines are essential to increase vaccine confidence among HCPs and patients.^34–36^

The clinical understanding of the viral mutations, at-risk population, long-term complications, and optimal vaccine schedule for COVID-19 is still evolving. Data to monitor trends in the dominant variant, test positivity rate, vaccination rates, hospitalization incidence, and characteristics of hospitalized patients exist but are located in disparate and sometimes inaccessible sources. The true utility of this data to impact clinical practice can only happen if the data is synthesized in one location with an intuitive user interface so that healthcare professionals can rapidly assess the current state of the pandemic.

There are several limitations to this analysis. First, there are risks of both overestimating the incidence of medically-attended COVID-19 and influenza. The use of diagnosis codes instead of positive laboratory tests risks overestimating the incidence of infection, as many respiratory infections have overlapping clinical presentations. However, during influenza season, this likely biases towards influenza being the preferred diagnosis. It may be biased towards COVID-19 if we are also capturing encounters for treatment of long COVID that are incorrectly coded for COVID-19. Previous research has shown low use of the diagnosis code specific to long COVID.^37^

The 6-month study period used in this analysis did not capture the full 2022-2023 influenza season as defined by the CDC. However, the peak influenza period is captured in this analysis, and the exclusion of the tail end of the season should not be expected to significantly change the results. This study took an inclusive approach to building code sets and, therefore, may be overestimating the prevalence of underlying medical conditions, including the use of immunosuppressive medications. The use of immunosuppressive medications found in this study, particularly among children, was very high but likely reflects the use of low doses of prednisone and topical corticosteroids, which are not anticipated to have strong immunosuppressive effects. The use of a broad approach biases towards no difference between cohorts, though it likely overestimates the number of immunosuppressed individuals.

The linked data source includes only insured individuals, and the results may not be representative of patients who are uninsured. In addition, we restricted to individuals with at least one medical record or claim between October 1, 2022, and March 31, 2023, so the population is likely to have a higher comorbidity burden than the US population as they represent people who are actively seeking care. Lastly, the pediatric population is underrepresented in the linked data source as compared to the US population. Therefore the pediatric infection burden might be underestimated when compared to older age cohorts.

## Conclusions

During the 2022-2023 influenza season, nearly three years after the start of the pandemic, medical encounters with a diagnosis of COVID-19 were more common than medical encounters with a diagnosis of influenza. In particular, hospitalizations with COVID-19 were more common than hospitalizations with influenza in all age subgroups examined.

## Data Availability

The data that support the findings of this study were used under license from Veradigm and Komodo Health. Due to data use agreements and its proprietary nature, restrictions apply regarding the availability of the data. Further information is available from the corresponding author.

## Funding

This work was supported by Moderna Inc.

## Disclosures/Conflicts of Interest

JAM and HK are employees of and shareholders in Moderna, Inc. AB, JW, CA, IW, and MB are employees of Veradigm, which was contracted by Moderna and received fees for data management and statistical analyses. VHN is an employee of VHN Consulting which was contracted by Moderna to help conduct this analysis.

## Acknowledgments

Ni Zeng, PhD, an employee of Veradigm, provided programming support and QA for this analysis. This support was funded by Moderna, Inc.

## Author Contributions

JM and HK contributed to the conceptualization of the study. AB, JW, CA and IW were responsible for data handling and formal analysis of the data. All authors contributed to the development of the methodology implemented in the study, data visualization, interpretation of the results, and writing and revising of the manuscript. All authors approved the final submitted version of the manuscript.

## Ethics approval

As a noninterventional, retrospective database study using a certified Health Insurance Portability and Accountability Act-compliant deidentified research database, approval by an institutional review board was not necessary.

