## Supplementary File 1 for "Comparison of COVID-19 and Influenza-Related Outcomes in the United States during Fall-Winter 2022-2023"

Supplementary File 1. Code lists

| **Condition** | **Type** | **Codes** |
| --- | --- | --- |
| Attention deficit and hyperactivity disorder (ADHD) | ICD-10-CM Dx | F90, F900, F901, F902, F908, F909 |
| Asthma | ICD-10-CM Dx | J45, J452, J4520, J4521, J4522, J453, J4530, J4531, J4532, J454, J4540, J4541, J4542, J455, J4550, J4551, J4552, J459, J45901, J45902, J45909, J45990, J45991, J45998 |
| Cancer | ICD-10-CM Dx | C000, C001, C002, C003, C004, C005, C006, C008, C009, C01, C020, C021, C022, C023, C024, C028, C029, C030, C031, C039, C040, C041, C048, C049, C050, C051, C052, C058, C059, C060, C061, C062, C0680, C0689, C069, C07, C080, C081, C089, C090, C091, C098, C099, C100, C101, C102, C103, C104, C108, C109, C110, C111, C112, C113, C118, C119, C12, C130, C131, C132, C138, C139, C140, C142, C148, C153, C154, C155, C158, C159, C160, C161, C162, C163, C164, C165, C166, C168, C169, C170, C171, C172, C173, C178, C179, C180, C181, C182, C183, C184, C185, C186, C187, C188, C189, C19, C20, C210, C211, C212, C218, C220, C221, C222, C223, C224, C227, C228, C229, C23, C240, C241, C248, C249, C250, C251, C252, C253, C254, C257, C258, C259, C260, C261, C269, C300, C301, C310, C311, C312, C313, C318, C319, C320, C321, C322, C323, C328, C329, C33, C3400, C3401, C3402, C3410, C3411, C3412, C342, C3430, C3431, C3432, C3480, C3481, C3482, C3490, C3491, C3492, C37, C380, C381, C382, C383, C384, C388, C390, C399, C4000, C4001, C4002, C4010, C4011, C4012, C4020, C4021, C4022, C4030, C4031, C4032, C4080, C4081, C4082, C4090, C4091, C4092, C410, C411, C412, C413, C414, C419, C430, C4310, C43111, C43112, C43121, C43122, C4320, C4321, C4322, C4330, C4331, C4339, C434, C4351, C4352, C4359, C4360, C4361, C4362, C4370, C4371, C4372, C438, C439, C4A0, C4A10, C4A111, C4A112, C4A121, C4A122, C4A20, C4A21, C4A22, C4A30, C4A31, C4A39, C4A4, C4A51, C4A52, C4A59, C4A60, C4A61, C4A62, C4A70, C4A71, C4A72, C4A8, C4A9, C4400, C4401, C4402, C4409, C44101, C441021, C441022, C441091, C441092, C44111, C441121, C441122, C441191, C441192, C44121, C441221, C441222, C441291, C441292, C44131, C441321, C441322, C441391, C441392, C44191, C441921, C441922, C441991, C441992, C44201, C44202, C44209, C44211, C44212, C44219, C44221, C44222, C44229, C44291, C44292, C44299, C44300, C44301, C44309, C44310, C44311, C44319, C44320, C44321, C44329, C44390, C44391, C44399, C4440, C4441, C4442, C4449, C44500, C44501, C44509, C44510, C44511, C44519, C44520, C44521, C44529, C44590, C44591, C44599, C44601, C44602, C44609, C44611, C44612, C44619, C44621, C44622, C44629, C44691, C44692, C44699, C44701, C44702, C44709, C44711, C44712, C44719, C44721, C44722, C44729, C44791, C44792, C44799, C4480, C4481, C4482, C4489, C4490, C4491, C4492, C4499, C450, C451, C452, C457, C459, C460, C461, C462, C463, C464, C4650, C4651, C4652, C467, C469, C470, C4710, C4711, C4712, C4720, C4721, C4722, C473, C474, C475, C476, C478, C479, C480, C481, C482, C488, C490, C4910, C4911, C4912, C4920, C4921, C4922, C493, C494, C495, C496, C498, C499, C49A0, C49A1, C49A2, C49A3, C49A4, C49A5, C49A9, C50011, C50012, C50019, C50021, C50022, C50029, C50111, C50112, C50119, C50121, C50122, C50129, C50211, C50212, C50219, C50221, C50222, C50229, C50311, C50312, C50319, C50321, C50322, C50329, C50411, C50412, C50419, C50421, C50422, C50429, C50511, C50512, C50519, C50521, C50522, C50529, C50611, C50612, C50619, C50621, C50622, C50629, C50811, C50812, C50819, C50821, C50822, C50829, C50911, C50912, C50919, C50921, C50922, C50929, C510, C511, C512, C518, C519, C52, C530, C531, C538, C539, C540, C541, C542, C543, C548, C549, C55, C561, C562, C569, C5700, C5701, C5702, C5710, C5711, C5712, C5720, C5721, C5722, C573, C574, C577, C578, C579, C58, C600, C601, C602, C608, C609, C61, C6200, C6201, C6202, C6210, C6211, C6212, C6290, C6291, C6292, C6300, C6301, C6302, C6310, C6311, C6312, C632, C637, C638, C639, C641, C642, C649, C651, C652, C659, C661, C662, C669, C670, C671, C672, C673, C674, C675, C676, C677, C678, C679, C680, C681, C688, C689, C6900, C6901, C6902, C6910, C6911, C6912, C6920, C6921, C6922, C6930, C6931, C6932, C6940, C6941, C6942, C6950, C6951, C6952, C6960, C6961, C6962, C6980, C6981, C6982, C6990, C6991, C6992, C700, C701, C709, C710, C711, C712, C713, C714, C715, C716, C717, C718, C719, C720, C721, C7220, C7221, C7222, C7230, C7231, C7232, C7240, C7241, C7242, C7250, C7259, C729, C73, C7400, C7401, C7402, C7410, C7411, C7412, C7490, C7491, C7492, C750, C751, C752, C753, C754, C755, C758, C759, C7A00, C7A010, C7A011, C7A012, C7A019, C7A020, C7A021, C7A022, C7A023, C7A024, C7A025, C7A026, C7A029, C7A090, C7A091, C7A092, C7A093, C7A094, C7A095, C7A096, C7A098, C7A1, C7A8, C7B00, C7B01, C7B02, C7B03, C7B04, C7B09, C7B1, C7B8, C760, C761, C762, C763, C7640, C7641, C7642, C7650, C7651, C7652, C768, C770, C771, C772, C773, C774, C775, C778, C779, C7800, C7801, C7802, C781, C782, C7830, C7839, C784, C785, C786, C787, C7880, C7889, C7900, C7901, C7902, C7910, C7911, C7919, C792, C7931, C7932, C7940, C7949, C7951, C7952, C7960, C7961, C7962, C7970, C7971, C7972, C7981, C7982, C7989, C799, C800, C801, C802, C8100, C8101, C8102, C8103, C8104, C8105, C8106, C8107, C8108, C8109, C8110, C8111, C8112, C8113, C8114, C8115, C8116, C8117, C8118, C8119, C8120, C8121, C8122, C8123, C8124, C8125, C8126, C8127, C8128, C8129, C8130, C8131, C8132, C8133, C8134, C8135, C8136, C8137, C8138, C8139, C8140, C8141, C8142, C8143, C8144, C8145, C8146, C8147, C8148, C8149, C8170, C8171, C8172, C8173, C8174, C8175, C8176, C8177, C8178, C8179, C8190, C8191, C8192, C8193, C8194, C8195, C8196, C8197, C8198, C8199, C8200, C8201, C8202, C8203, C8204, C8205, C8206, C8207, C8208, C8209, C8210, C8211, C8212, C8213, C8214, C8215, C8216, C8217, C8218, C8219, C8220, C8221, C8222, C8223, C8224, C8225, C8226, C8227, C8228, C8229, C8230, C8231, C8232, C8233, C8234, C8235, C8236, C8237, C8238, C8239, C8240, C8241, C8242, C8243, C8244, C8245, C8246, C8247, C8248, C8249, C8250, C8251, C8252, C8253, C8254, C8255, C8256, C8257, C8258, C8259, C8260, C8261, C8262, C8263, C8264, C8265, C8266, C8267, C8268, C8269, C8280, C8281, C8282, C8283, C8284, C8285, C8286, C8287, C8288, C8289, C8290, C8291, C8292, C8293, C8294, C8295, C8296, C8297, C8298, C8299, C8300, C8301, C8302, C8303, C8304, C8305, C8306, C8307, C8308, C8309, C8310, C8311, C8312, C8313, C8314, C8315, C8316, C8317, C8318, C8319, C8330, C8331, C8332, C8333, C8334, C8335, C8336, C8337, C8338, C8339, C8350, C8351, C8352, C8353, C8354, C8355, C8356, C8357, C8358, C8359, C8370, C8371, C8372, C8373, C8374, C8375, C8376, C8377, C8378, C8379, C8380, C8381, C8382, C8383, C8384, C8385, C8386, C8387, C8388, C8389, C8390, C8391, C8392, C8393, C8394, C8395, C8396, C8397, C8398, C8399, C8400, C8401, C8402, C8403, C8404, C8405, C8406, C8407, C8408, C8409, C8410, C8411, C8412, C8413, C8414, C8415, C8416, C8417, C8418, C8419, C8440, C8441, C8442, C8443, C8444, C8445, C8446, C8447, C8448, C8449, C8460, C8461, C8462, C8463, C8464, C8465, C8466, C8467, C8468, C8469, C8470, C8471, C8472, C8473, C8474, C8475, C8476, C8477, C8478, C8479, C84A0, C84A1, C84A2, C84A3, C84A4, C84A5, C84A6, C84A7, C84A8, C84A9, C84Z0, C84Z1, C84Z2, C84Z3, C84Z4, C84Z5, C84Z6, C84Z7, C84Z8, C84Z9, C8490, C8491, C8492, C8493, C8494, C8495, C8496, C8497, C8498, C8499, C8510, C8511, C8512, C8513, C8514, C8515, C8516, C8517, C8518, C8519, C8520, C8521, C8522, C8523, C8524, C8525, C8526, C8527, C8528, C8529, C8580, C8581, C8582, C8583, C8584, C8585, C8586, C8587, C8588, C8589, C8590, C8591, C8592, C8593, C8594, C8595, C8596, C8597, C8598, C8599, C860, C861, C862, C863, C864, C865, C866, C880, C882, C883, C884, C888, C889, C9000, C9001, C9002, C9010, C9011, C9012, C9020, C9021, C9022, C9030, C9031, C9032, C9100, C9101, C9102, C9110, C9111, C9112, C9130, C9131, C9132, C9140, C9141, C9142, C9150, C9151, C9152, C9160, C9161, C9162, C91A0, C91A1, C91A2, C91Z0, C91Z1, C91Z2, C9190, C9191, C9192, C9200, C9201, C9202, C9210, C9211, C9212, C9220, C9221, C9222, C9230, C9231, C9232, C9240, C9241, C9242, C9250, C9251, C9252, C9260, C9261, C9262, C92A0, C92A1, C92A2, C92Z0, C92Z1, C92Z2, C9290, C9291, C9292, C9300, C9301, C9302, C9310, C9311, C9312, C9330, C9331, C9332, C93Z0, C93Z1, C93Z2, C9390, C9391, C9392, C9400, C9401, C9402, C9420, C9421, C9422, C9430, C9431, C9432, C9440, C9441, C9442, C946, C9480, C9481, C9482, C9500, C9501, C9502, C9510, C9511, C9512, C9590, C9591, C9592, C960, C9620, C9621, C9622, C9629, C964, C965, C966, C96A, C96Z, C969, D0000, D0001, D0002, D0003, D0004, D0005, D0006, D0007, D0008, D001, D002, D010, D011, D012, D013, D0140, D0149, D015, D017, D019, D020, D021, D0220, D0221, D0222, D023, D024, D030, D0310, D03111, D03112, D03121, D03122, D0320, D0321, D0322, D0330, D0339, D034, D0351, D0352, D0359, D0360, D0361, D0362, D0370, D0371, D0372, D038, D039, D040, D0410, D04111, D04112, D04121, D04122, D0420, D0421, D0422, D0430, D0439, D044, D045, D0460, D0461, D0462, D0470, D0471, D0472, D048, D049, D0500, D0501, D0502, D0510, D0511, D0512, D0580, D0581, D0582, D0590, D0591, D0592, D060, D061, D067, D069, D070, D071, D072, D0730, D0739, D074, D075, D0760, D0761, D0769, D090, D0910, D0919, D0920, D0921, D0922, D093, D098, D099, D100, D101, D102, D1030, D1039, D104, D105, D106, D107, D109, D110, D117, D119, D120, D121, D122, D123, D124, D125, D126, D127, D128, D129, D130, D131, D132, D1330, D1339, D134, D135, D136, D137, D139, D140, D141, D142, D1430, D1431, D1432, D144, D150, D151, D152, D157, D159, D1600, D1601, D1602, D1610, D1611, D1612, D1620, D1621, D1622, D1630, D1631, D1632, D164, D165, D166, D167, D168, D169, D170, D171, D1720, D1721, D1722, D1723, D1724, D1730, D1739, D174, D175, D176, D1771, D1772, D1779, D179, D1800, D1801, D1802, D1803, D1809, D181, D190, D191, D197, D199, D200, D201, D210, D2110, D2111, D2112, D2120, D2121, D2122, D213, D214, D215, D216, D219, D220, D2210, D22111, D22112, D22121, D22122, D2220, D2221, D2222, D2230, D2239, D224, D225, D2260, D2261, D2262, D2270, D2271, D2272, D229, D230, D2310, D23111, D23112, D23121, D23122, D2320, D2321, D2322, D2330, D2339, D234, D235, D2360, D2361, D2362, D2370, D2371, D2372, D239, D241, D242, D249, D250, D251, D252, D259, D260, D261, D267, D269, D270, D271, D279, D280, D281, D282, D287, D289, D290, D291, D2920, D2921, D2922, D2930, D2931, D2932, D294, D298, D299, D3000, D3001, D3002, D3010, D3011, D3012, D3020, D3021, D3022, D303, D304, D308, D309, D3100, D3101, D3102, D3110, D3111, D3112, D3120, D3121, D3122, D3130, D3131, D3132, D3140, D3141, D3142, D3150, D3151, D3152, D3160, D3161, D3162, D3190, D3191, D3192, D320, D321, D329, D330, D331, D332, D333, D334, D337, D339, D34, D3500, D3501, D3502, D351, D352, D353, D354, D355, D356, D357, D359, D360, D3610, D3611, D3612, D3613, D3614, D3615, D3616, D3617, D367, D369, D3A00, D3A010, D3A011, D3A012, D3A019, D3A020, D3A021, D3A022, D3A023, D3A024, D3A025, D3A026, D3A029, D3A090, D3A091, D3A092, D3A093, D3A094, D3A095, D3A096, D3A098, D3A8, D3701, D3702, D37030, D37031, D37032, D37039, D3704, D3705, D3709, D371, D372, D373, D374, D375, D376, D378, D379, D380, D381, D382, D383, D384, D385, D386, D390, D3910, D3911, D3912, D392, D398, D399, D400, D4010, D4011, D4012, D408, D409, D4100, D4101, D4102, D4110, D4111, D4112, D4120, D4121, D4122, D413, D414, D418, D419, D420, D421, D429, D430, D431, D432, D433, D434, D438, D439, D440, D4410, D4411, D4412, D442, D443, D444, D445, D446, D447, D449, D45, D460, D461, D4620, D4621, D4622, D46A, D46B, D46C, D464, D46Z, D469, D4701, D4702, D4709, D471, D472, D473, D474, D47Z1, D47Z2, D47Z9, D479, D480, D481, D482, D483, D484, D485, D4860, D4861, D4862, D487, D489, D490, D491, D492, D493, D494, D49511, D49512, D49519, D4959, D496, D497, D4981, D4989, D499 |
| Cerebral palsy | ICD-10-CM Dx | G80, G800, G801, G802, G803, G804, G808, G809 |
| Cerebrovascular disease | ICD-10-CM Dx | I6000, I6001, I6002, I6010, I6011, I6012, I602, I6030, I6031, I6032, I604, I6050, I6051, I6052, I606, I607, I608, I609, I610, I611, I612, I613, I614, I615, I616, I618, I619, I6200, I6201, I6202, I6203, I621, I629, I6300, I63011, I63012, I63013, I63019, I6302, I63031, I63032, I63033, I63039, I6309, I6310, I63111, I63112, I63113, I63119, I6312, I63131, I63132, I63133, I63139, I6319, I6320, I63211, I63212, I63213, I63219, I6322, I63231, I63232, I63233, I63239, I6329, I6330, I63311, I63312, I63313, I63319, I63321, I63322, I63323, I63329, I63331, I63332, I63333, I63339, I63341, I63342, I63343, I63349, I6339, I6340, I63411, I63412, I63413, I63419, I63421, I63422, I63423, I63429, I63431, I63432, I63433, I63439, I63441, I63442, I63443, I63449, I6349, I6350, I63511, I63512, I63513, I63519, I63521, I63522, I63523, I63529, I63531, I63532, I63533, I63539, I63541, I63542, I63543, I63549, I6359, I636, I6381, I6389, I639, I6501, I6502, I6503, I6509, I651, I6521, I6522, I6523, I6529, I658, I659, I6601, I6602, I6603, I6609, I6611, I6612, I6613, I6619, I6621, I6622, I6623, I6629, I663, I668, I669, I670, I671, I672, I673, I674, I675, I676, I677, I6781, I6782, I6783, I67841, I67848, I67850, I67858, I6789, I679, I680, I682, I688, I6900, I69010, I69011, I69012, I69013, I69014, I69015, I69018, I69019, I69020, I69021, I69022, I69023, I69028, I69031, I69032, I69033, I69034, I69039, I69041, I69042, I69043, I69044, I69049, I69051, I69052, I69053, I69054, I69059, I69061, I69062, I69063, I69064, I69065, I69069, I69090, I69091, I69092, I69093, I69098, I6910, I69110, I69111, I69112, I69113, I69114, I69115, I69118, I69119, I69120, I69121, I69122, I69123, I69128, I69131, I69132, I69133, I69134, I69139, I69141, I69142, I69143, I69144, I69149, I69151, I69152, I69153, I69154, I69159, I69161, I69162, I69163, I69164, I69165, I69169, I69190, I69191, I69192, I69193, I69198, I6920, I69210, I69211, I69212, I69213, I69214, I69215, I69218, I69219, I69220, I69221, I69222, I69223, I69228, I69231, I69232, I69233, I69234, I69239, I69241, I69242, I69243, I69244, I69249, I69251, I69252, I69253, I69254, I69259, I69261, I69262, I69263, I69264, I69265, I69269, I69290, I69291, I69292, I69293, I69298, I6930, I69310, I69311, I69312, I69313, I69314, I69315, I69318, I69319, I69320, I69321, I69322, I69323, I69328, I69331, I69332, I69333, I69334, I69339, I69341, I69342, I69343, I69344, I69349, I69351, I69352, I69353, I69354, I69359, I69361, I69362, I69363, I69364, I69365, I69369, I69390, I69391, I69392, I69393, I69398, I6980, I69810, I69811, I69812, I69813, I69814, I69815, I69818, I69819, I69820, I69821, I69822, I69823, I69828, I69831, I69832, I69833, I69834, I69839, I69841, I69842, I69843, I69844, I69849, I69851, I69852, I69853, I69854, I69859, I69861, I69862, I69863, I69864, I69865, I69869, I69890, I69891, I69892, I69893, I69898, I6990, I69910, I69911, I69912, I69913, I69914, I69915, I69918, I69919, I69920, I69921, I69922, I69923, I69928, I69931, I69932, I69933, I69934, I69939, I69941, I69942, I69943, I69944, I69949, I69951, I69952, I69953, I69954, I69959, I69961, I69962, I69963, I69964, I69965, I69969, I69990, I69991, I69992, I69993, I69998 |
| Chronic kidney disease | ICD-10-CM Dx | N181, N182, N1830, N1831, N1832, N184, N185, N186, N189 |
| Chronic liver disease | ICD-10-CM Dx | K743, K744, K745, K7460, K7469, K7581, K7589, K754, K700, K7010, K7011, K702, K7030, K7031, K7040, K7041, K709 |
| Chronic lung disease | ICD-10-CM Dx | J80, J810, J811, J8281, J8282, J8283, J8289, J8401, J8402, J8403, J8409, J8410, J84111, J84112, J84113, J84114, J84115, J84116, J84117, J84170, J84178, J842, J8481, J8482, J8483, J84841, J84842, J84843, J84848, J8489, J849, I2601, I2602, I2609, I2690, I2692, I2693, I2694, I2699, I2720, I2721, I2722, I2723, I2724, I2729, J470, J471, J479, J440, J441, J449, P271 |
| Congenital malformation | ICD-10-CM Dx | Q00, Q000, Q001, Q002, Q01, Q010, Q011, Q012, Q018, Q019, Q02, Q03, Q030, Q031, Q038, Q039, Q04, Q040, Q041, Q042, Q043, Q044, Q045, Q046, Q048, Q049, Q05, Q050, Q051, Q052, Q053, Q054, Q055, Q056, Q057, Q058, Q059, Q06, Q060, Q061, Q062, Q063, Q064, Q068, Q069, Q07, Q070, Q0700, Q0701, Q0702, Q0703, Q078, Q079, Q10, Q100, Q101, Q102, Q103, Q104, Q105, Q106, Q107, Q11, Q110, Q111, Q112, Q113, Q12, Q120, Q121, Q122, Q123, Q124, Q128, Q129, Q13, Q130, Q131, Q132, Q133, Q134, Q135, Q138, Q1381, Q1389, Q139, Q14, Q140, Q141, Q142, Q143, Q148, Q149, Q15, Q150, Q158, Q159, Q16, Q160, Q161, Q162, Q163, Q164, Q165, Q169, Q17, Q170, Q171, Q172, Q173, Q174, Q175, Q178, Q179, Q18, Q180, Q181, Q182, Q183, Q184, Q185, Q186, Q187, Q188, Q189, Q20, Q200, Q201, Q202, Q203, Q204, Q205, Q206, Q208, Q209, Q21, Q210, Q211, Q212, Q213, Q214, Q218, Q219, Q22, Q220, Q221, Q222, Q223, Q224, Q225, Q226, Q228, Q229, Q23, Q230, Q231, Q232, Q233, Q234, Q238, Q239, Q24, Q240, Q241, Q242, Q243, Q244, Q245, Q246, Q248, Q249, Q25, Q250, Q251, Q252, Q2521, Q2529, Q253, Q254, Q2540, Q2541, Q2542, Q2543, Q2544, Q2545, Q2546, Q2547, Q2548, Q2549, Q255, Q256, Q257, Q2571, Q2572, Q2579, Q258, Q259, Q26, Q260, Q261, Q262, Q263, Q264, Q265, Q266, Q268, Q269, Q27, Q270, Q271, Q272, Q273, Q2730, Q2731, Q2732, Q2733, Q2734, Q2739, Q274, Q278, Q279, Q28, Q280, Q281, Q282, Q283, Q288, Q289, Q30, Q300, Q301, Q302, Q303, Q308, Q309, Q31, Q310, Q311, Q312, Q313, Q315, Q318, Q319, Q32, Q320, Q321, Q322, Q323, Q324, Q33, Q330, Q331, Q332, Q333, Q334, Q335, Q336, Q338, Q339, Q34, Q340, Q341, Q348, Q349, Q35, Q351, Q353, Q355, Q357, Q359, Q36, Q360, Q361, Q369, Q37, Q370, Q371, Q372, Q373, Q374, Q375, Q378, Q379, Q38, Q380, Q381, Q382, Q383, Q384, Q385, Q386, Q387, Q388, Q39, Q390, Q391, Q392, Q393, Q394, Q395, Q396, Q398, Q399, Q40, Q400, Q401, Q402, Q403, Q408, Q409, Q41, Q410, Q411, Q412, Q418, Q419, Q42, Q420, Q421, Q422, Q423, Q428, Q429, Q43, Q430, Q431, Q432, Q433, Q434, Q435, Q436, Q437, Q438, Q439, Q44, Q440, Q441, Q442, Q443, Q444, Q445, Q446, Q447, Q45, Q450, Q451, Q452, Q453, Q458, Q459, Q50, Q500, Q5001, Q5002, Q501, Q502, Q503, Q5031, Q5032, Q5039, Q504, Q505, Q506, Q51, Q510, Q511, Q5110, Q5111, Q512, Q5121, Q5122, Q5128, Q513, Q514, Q515, Q516, Q517, Q518, Q5181, Q51810, Q51811, Q51818, Q5182, Q51820, Q51821, Q51828, Q519, Q52, Q520, Q521, Q5210, Q5211, Q5212, Q52120, Q52121, Q52122, Q52123, Q52124, Q52129, Q522, Q523, Q524, Q525, Q526, Q527, Q5270, Q5271, Q5279, Q528, Q529, Q53, Q530, Q5300, Q5301, Q5302, Q531, Q5310, Q5311, Q53111, Q53112, Q5312, Q5313, Q532, Q5320, Q5321, Q53211, Q53212, Q5322, Q5323, Q539, Q54, Q540, Q541, Q542, Q543, Q544, Q548, Q549, Q55, Q550, Q551, Q552, Q5520, Q5521, Q5522, Q5523, Q5529, Q553, Q554, Q555, Q556, Q5561, Q5562, Q5563, Q5564, Q5569, Q557, Q558, Q559, Q56, Q560, Q561, Q562, Q563, Q564, Q60, Q600, Q601, Q602, Q603, Q604, Q605, Q606, Q61, Q610, Q6100, Q6101, Q6102, Q611, Q6111, Q6119, Q612, Q613, Q614, Q615, Q618, Q619, Q62, Q620, Q621, Q6210, Q6211, Q6212, Q622, Q623, Q6231, Q6232, Q6239, Q624, Q625, Q626, Q6260, Q6261, Q6262, Q6263, Q6269, Q627, Q628, Q63, Q630, Q631, Q632, Q633, Q638, Q639, Q64, Q640, Q641, Q6410, Q6411, Q6412, Q6419, Q642, Q643, Q6431, Q6432, Q6433, Q6439, Q644, Q645, Q646, Q647, Q6470, Q6471, Q6472, Q6473, Q6474, Q6475, Q6479, Q648, Q649, Q65, Q650, Q6500, Q6501, Q6502, Q651, Q652, Q653, Q6530, Q6531, Q6532, Q654, Q655, Q656, Q658, Q6581, Q6582, Q6589, Q659, Q66, Q660, Q6600, Q6601, Q6602, Q661, Q6610, Q6611, Q6612, Q662, Q6621, Q66211, Q66212, Q66219, Q6622, Q66221, Q66222, Q66229, Q663, Q6630, Q6631, Q6632, Q664, Q6640, Q6641, Q6642, Q665, Q6650, Q6651, Q6652, Q666, Q667, Q6670, Q6671, Q6672, Q668, Q6680, Q6681, Q6682, Q6689, Q669, Q6690, Q6691, Q6692, Q67, Q670, Q671, Q672, Q673, Q674, Q675, Q676, Q677, Q678, Q68, Q680, Q681, Q682, Q683, Q684, Q685, Q686, Q688, Q69, Q690, Q691, Q692, Q699, Q70, Q700, Q7000, Q7001, Q7002, Q7003, Q701, Q7010, Q7011, Q7012, Q7013, Q702, Q7020, Q7021, Q7022, Q7023, Q703, Q7030, Q7031, Q7032, Q7033, Q704, Q709, Q71, Q710, Q7100, Q7101, Q7102, Q7103, Q711, Q7110, Q7111, Q7112, Q7113, Q712, Q7120, Q7121, Q7122, Q7123, Q713, Q7130, Q7131, Q7132, Q7133, Q714, Q7140, Q7141, Q7142, Q7143, Q715, Q7150, Q7151, Q7152, Q7153, Q716, Q7160, Q7161, Q7162, Q7163, Q718, Q7181, Q71811, Q71812, Q71813, Q71819, Q7189, Q71891, Q71892, Q71893, Q71899, Q719, Q7190, Q7191, Q7192, Q7193, Q72, Q720, Q7200, Q7201, Q7202, Q7203, Q721, Q7210, Q7211, Q7212, Q7213, Q722, Q7220, Q7221, Q7222, Q7223, Q723, Q7230, Q7231, Q7232, Q7233, Q724, Q7240, Q7241, Q7242, Q7243, Q725, Q7250, Q7251, Q7252, Q7253, Q726, Q7260, Q7261, Q7262, Q7263, Q727, Q7270, Q7271, Q7272, Q7273, Q728, Q7281, Q72811, Q72812, Q72813, Q72819, Q7289, Q72891, Q72892, Q72893, Q72899, Q729, Q7290, Q7291, Q7292, Q7293, Q73, Q730, Q731, Q738, Q74, Q740, Q741, Q742, Q743, Q748, Q749, Q75, Q750, Q751, Q752, Q753, Q754, Q755, Q758, Q759, Q76, Q760, Q761, Q762, Q763, Q764, Q7641, Q76411, Q76412, Q76413, Q76414, Q76415, Q76419, Q7642, Q76425, Q76426, Q76427, Q76428, Q76429, Q7649, Q765, Q766, Q767, Q768, Q769, Q77, Q770, Q771, Q772, Q773, Q774, Q775, Q776, Q777, Q778, Q779, Q78, Q780, Q781, Q782, Q783, Q784, Q785, Q786, Q788, Q789, Q79, Q790, Q791, Q792, Q793, Q794, Q795, Q7951, Q7959, Q796, Q7960, Q7961, Q7962, Q7963, Q7969, Q798, Q799, Q80, Q800, Q801, Q802, Q803, Q804, Q808, Q809, Q81, Q810, Q811, Q812, Q818, Q819, Q82, Q820, Q821, Q822, Q823, Q824, Q825, Q826, Q828, Q829, Q83, Q830, Q831, Q832, Q833, Q838, Q839, Q84, Q840, Q841, Q842, Q843, Q844, Q845, Q846, Q848, Q849, Q85, Q850, Q8500, Q8501, Q8502, Q8503, Q8509, Q851, Q858, Q859, Q86, Q860, Q861, Q862, Q868, Q87, Q870, Q871, Q8711, Q8719, Q872, Q873, Q874, Q8740, Q8741, Q87410, Q87418, Q8742, Q8743, Q875, Q878, Q8781, Q8782, Q8789, Q89, Q890, Q8901, Q8909, Q891, Q892, Q893, Q894, Q897, Q898, Q899 |
| COVID-19 | ICD-10-CM Dx | U071, B342, B9729, Z8616, J1282 |
|  | ICD10 | U072 |
|  | SNOMED | 840533007, 840539006, 1119302008, 119731000146105, 119741000146102, 119751000146104, 119981000146107, 1240521000000100, 1240531000000100, 1240541000000100, 1240561000000100, 1240581000000100, 674814021000119000, 866151004, 866152006, 870577009, 870588003, 870589006, 870590002, 870591003, 871562009, 1240411000000100, 840536004, 840534001 |
| Cystic fibrosis | ICD-10-CM Dx | E84, E840, E841, E8411, E8419, E848, E849 |
| Dementia (only in individuals 18+) | ICD-10-CM Dx | F01, F015, F0150, F0151, F02, F028, F0280, F0281, F03, F039, F0390, F0391, F04, F05, F061, F068, G132, G138, G30, G300, G301, G308, G309, G310, G3101, G3109, G311, G312, G914, G94, R4181, R54 |
| Diabetes (type 1 and type 2) | ICD-10-CM Dx | E1010, E1011, E1021, E1022, E1029, E10311, E10319, E103211, E103212, E103213, E103219, E103291, E103292, E103293, E103299, E103311, E103312, E103313, E103319, E103391, E103392, E103393, E103399, E103411, E103412, E103413, E103419, E103491, E103492, E103493, E103499, E103511, E103512, E103513, E103519, E103521, E103522, E103523, E103529, E103531, E103532, E103533, E103539, E103541, E103542, E103543, E103549, E103551, E103552, E103553, E103559, E103591, E103592, E103593, E103599, E1036, E1037X1, E1037X2, E1037X3, E1037X9, E1039, E1040, E1041, E1042, E1043, E1044, E1049, E1051, E1052, E1059, E10610, E10618, E10620, E10621, E10622, E10628, E10630, E10638, E10641, E10649, E1065, E1069, E108, E109, E1100, E1101, E1110, E1111, E1121, E1122, E1129, E11311, E11319, E113211, E113212, E113213, E113219, E113291, E113292, E113293, E113299, E113311, E113312, E113313, E113319, E113391, E113392, E113393, E113399, E113411, E113412, E113413, E113419, E113491, E113492, E113493, E113499, E113511, E113512, E113513, E113519, E113521, E113522, E113523, E113529, E113531, E113532, E113533, E113539, E113541, E113542, E113543, E113549, E113551, E113552, E113553, E113559, E113591, E113592, E113593, E113599, E1136, E1137X1, E1137X2, E1137X3, E1137X9, E1139, E1140, E1141, E1142, E1143, E1144, E1149, E1151, E1152, E1159, E11610, E11618, E11620, E11621, E11622, E11628, E11630, E11638, E11641, E11649, E1165, E1169, E118, E119 |
| Disability | ICD-10-CM Dx | F70, F71, F72, F73, F78, F78A, F78A1, F78A9, F79, F80, F800, F801, F802, F804, F808, F8081, F8082, F8089, F809, F81, F810, F812, F818, F8181, F8189, F819, F82, F84, F840, F842, F843, F845, F848, F849, F88, F89 |
| Down syndrome | ICD-10-CM Dx | Q900, Q901, Q902, Q909 |
| Heart disease | ICD-10-CM Dx | I200, I201, I208, I209, I2101, I2102, I2109, I2111, I2119, I2121, I2129, I213, I214, I219, I21A1, I21A9, I220, I221, I222, I228, I229, I230, I231, I232, I233, I234, I235, I236, I237, I238, I240, I241, I248, I249, I2510, I25110, I25111, I25118, I25119, I252, I253, I2541, I2542, I255, I256, I25700, I25701, I25708, I25709, I25710, I25711, I25718, I25719, I25720, I25721, I25728, I25729, I25730, I25731, I25738, I25739, I25750, I25751, I25758, I25759, I25760, I25761, I25768, I25769, I25790, I25791, I25798, I25799, I25810, I25811, I25812, I2582, I2583, I2584, I2589, I259, I420, I421, I422, I423, I424, I425, I426, I427, I428, I429, I501, I5020, I5021, I5022, I5023, I5030, I5031, I5032, I5033, I5040, I5041, I5042, I5043, I50810, I50811, I50812, I50813, I50814, I5082, I5083, I5084, I5089, I509 |
| Human immunodeficiency virus (HIV) | ICD-10-CM Dx | B20, B9735, Z21 |
| Hypertension | ICD-10-CM Dx | H3503, H35031, H35032, H35033, H35039, I10, I11, I110, I119, I12, I120, I129, I13, I130, I131, I1310, I1311, I132, I15, I150, I151, I152, I158, I159, N262 |
| Influenza | ICD-10-CM Dx | J09, J09x, J09X1, J09X2, J09X3, J09X9, J10, J100, J1000, J1001, J1008, J101, J102, J108, J1081, J1082, J1083, J1089, J11, J110, J1100, J1108, J111, J112, J118, J1181, J1182, J1183, J1189 |
| Use of immunosuppressive medications (including topical corticosteroids) | CPT | 80158, 80197, 80169, 80180, 80195 |
|  | HCPCS | C9006, C9020, C9026, C9106, C9110, C9211, C9212, C9230, C9236, C9239, C9249, C9261, C9421, C9455, J0129, J0135, J0202, J0480, J0490, J0638, J1300, J1602, J1628, J2323, J2793, J3245, J3357, J7504, J7507, J7508, J7513, J7515, J7517, J7520, J8530, J9092, J9094, J9095, J9250, J9260, J9310, J9312, J9330, K0120, K0121, K0412, Q2019, Q2044, Q4079, Q5109, S0087, S0162, S0193, S9359, C9126, C9219, C9264, C9286, C9419, C9420, C9436, C9438, J0215, J0485, J0717, J0718, J1438, J1745, J2350, J2860, J3262, J3358, J3380, J7500, J7501, J7502, J7503, J7505, J7511, J7518, J7525, J7527, J8561, J8610, J9010, J9065, J9070, J9080, J9090, J9091, J9093, J9096, J9097, J9311, K0119, K0122, K0123, Q5103, Q5104 |
|  | NDC Generic Names | Abatacept, Adalimumab, Alclometasone, Alefacept, Alemtuzumab, Amcinonide, Anakinra, Apremilast, Azacitidine, Azathioprine, Baricitinib, Basiliximab, Beclomethasone, Belatacept, Belimumab, Bendamustine, Betamethasone, Brodalumab, Budesonide, Busulfan, Canakinumab, Capecitabine, Carmustine, Certolizumab Pegol, Chlorambucil, Ciclesonide, Cladribine, Clobetasol, Clofarabine, Cortisone, Cyclophosphamide, Cyclosporine, Cytarabine, Dacarbazine, Daclizumab, Daunorubicin-Cytarabine, Decitabine, Decitabine-Cedazuridine, Deflazacort, Desonide, Desoximetasone, Dexamethasone, Diflorasone, Difluprednate, Dimethyl Fumarate, Diroximel Fumarate, Eculizumab, Efalizumab, Emapalumab-lzsg, Etanercept, Everolimus, Fingolimod, Floxuridine, Fludarabine, Flunisolide, Fluocinolone, Fluocinonide, Fluorometholone, Fluorouracil, Flurandrenolide, Fluticasone, Gemcitabine, Golimumab, Guselkumab, Halcinonide, Halobetasol, Hydrocortisone, Ifosfamide, Ifosfamide-Mesna, Inebilizumab-cdon, Infliximab, Infliximab-abda, Infliximab-axxq, Infliximab-dyyb, Ixekizumab, Leflunomide, Lenalidomide, Lomustine, Loteprednol, Medrysone, Melphalan, Mercaptopurine, Methotrexate, Methylprednisolone, Mifepristone, Mometasone, Muromonab-Cd3 1, Mycophenolate Mofetil, Natalizumab, Nelarabine, Ocrelizumab, Ofatumumab, Ozanimod Hydrochloride, Pemetrexed Disodium, Pirfenidone, Pomalidomide, Pralatrexate, Prednicarbate, Prednisolone, Prednisone, Ravulizumab-cwvz, Rilonacept, Rimexolone, Risankizumab-rzaa, Sarilumab, Satralizumab-mwge, Secukinumab , Siltuximab, Siponimod, Sirolimus, Streptozocin, Tacrolimus, Temozolomide, Temsirolimus, Teprotumumab-trbw, Teriflunomide, Thalidomide, Thiotepa, Tildrakizumab-asmn, Tocilizumab, Tofacitinib, Triamcinolone, Triamcinolone , Upadacitinib, Ustekinumab, Vedolizumab |
| Mental health conditions | ICD-10-CM Dx | F200, F201, F202, F203, F205, F2081, F2089, F209, F21, F22, F23, F24, F250, F251, F258, F259, F28, F29, F3010, F3011, F3012, F3013, F302, F303, F304, F308, F309, F310, F3110, F3111, F3112, F3113, F312, F3130, F3131, F3132, F314, F315, F3160, F3161, F3162, F3163, F3164, F3170, F3171, F3172, F3173, F3174, F3175, F3176, F3177, F3178, F3181, F3189, F319, F320, F321, F322, F323, F324, F325, F3281, F3289, F329, F330, F331, F332, F333, F3340, F3341, F3342, F338, F339, F340, F341, F3481, F3489, F349, F39 |
| Musculoskeletal conditions | ICD-10-CM Dx | M050, M0500, M0501, M05011, M05012, M05019, M0502, M05021, M05022, M05029, M0503, M05031, M05032, M05039, M0504, M05041, M05042, M05049, M0505, M05051, M05052, M05059, M0506, M05061, M05062, M05069, M0507, M05071, M05072, M05079, M0509, M051, M0510, M0511, M05111, M05112, M05119, M0512, M05121, M05122, M05129, M0513, M05131, M05132, M05139, M0514, M05141, M05142, M05149, M0515, M05151, M05152, M05159, M0516, M05161, M05162, M05169, M0517, M05171, M05172, M05179, M0519, M052, M0520, M0521, M05211, M05212, M05219, M0522, M05221, M05222, M05229, M0523, M05231, M05232, M05239, M0524, M05241, M05242, M05249, M0525, M05251, M05252, M05259, M0526, M05261, M05262, M05269, M0527, M05271, M05272, M05279, M0529, M053, M0530, M0531, M05311, M05312, M05319, M0532, M05321, M05322, M05329, M0533, M05331, M05332, M05339, M0534, M05341, M05342, M05349, M0535, M05351, M05352, M05359, M0536, M05361, M05362, M05369, M0537, M05371, M05372, M05379, M0539, M054, M0540, M0541, M05411, M05412, M05419, M0542, M05421, M05422, M05429, M0543, M05431, M05432, M05439, M0544, M05441, M05442, M05449, M0545, M05451, M05452, M05459, M0546, M05461, M05462, M05469, M0547, M05471, M05472, M05479, M0549, M055, M0550, M0551, M05511, M05512, M05519, M0552, M05521, M05522, M05529, M0553, M05531, M05532, M05539, M0554, M05541, M05542, M05549, M0555, M05551, M05552, M05559, M0556, M05561, M05562, M05569, M0557, M05571, M05572, M05579, M0559, M056, M0560, M0561, M05611, M05612, M05619, M0562, M05621, M05622, M05629, M0563, M05631, M05632, M05639, M0564, M05641, M05642, M05649, M0565, M05651, M05652, M05659, M0566, M05661, M05662, M05669, M0567, M05671, M05672, M05679, M0569, M057, M0570, M0571, M05711, M05712, M05719, M0572, M05721, M05722, M05729, M0573, M05731, M05732, M05739, M0574, M05741, M05742, M05749, M0575, M05751, M05752, M05759, M0576, M05761, M05762, M05769, M0577, M05771, M05772, M05779, M0579, M057A, M058, M0580, M0581, M05811, M05812, M05819, M0582, M05821, M05822, M05829, M0583, M05831, M05832, M05839, M0584, M05841, M05842, M05849, M0585, M05851, M05852, M05859, M0586, M05861, M05862, M05869, M0587, M05871, M05872, M05879, M0589, M058A, M059, M06, M060, M0600, M0601, M06011, M06012, M06019, M0602, M06021, M06022, M06029, M0603, M06031, M06032, M06039, M0604, M06041, M06042, M06049, M0605, M06051, M06052, M06059, M0606, M06061, M06062, M06069, M0607, M06071, M06072, M06079, M0608, M0609, M060A, M061, M062, M0620, M0621, M06211, M06212, M06219, M0622, M06221, M06222, M06229, M0623, M06231, M06232, M06239, M0624, M06241, M06242, M06249, M0625, M06251, M06252, M06259, M0626, M06261, M06262, M06269, M0627, M06271, M06272, M06279, M0628, M0629, M063, M0630, M0631, M06311, M06312, M06319, M0632, M06321, M06322, M06329, M0633, M06331, M06332, M06339, M0634, M06341, M06342, M06349, M0635, M06351, M06352, M06359, M0636, M06361, M06362, M06369, M0637, M06371, M06372, M06379, M0638, M0639, M064, M068, M0680, M0681, M06811, M06812, M06819, M0682, M06821, M06822, M06829, M0683, M06831, M06832, M06839, M0684, M06841, M06842, M06849, M0685, M06851, M06852, M06859, M0686, M06861, M06862, M06869, M0687, M06871, M06872, M06879, M0688, M0689, M068A, M069, M07, M076, M0760, M0761, M07611, M07612, M07619, M0762, M07621, M07622, M07629, M0763, M07631, M07632, M07639, M0764, M07641, M07642, M07649, M0765, M07651, M07652, M07659, M0766, M07661, M07662, M07669, M0767, M07671, M07672, M07679, M0768, M0769, M08, M080, M0800, M0801, M08011, M08012, M08019, M0802, M08021, M08022, M08029, M0803, M08031, M08032, M08039, M0804, M08041, M08042, M08049, M0805, M08051, M08052, M08059, M0806, M08061, M08062, M08069, M0807, M08071, M08072, M08079, M0808, M0809, M080A, M081, M082, M0820, M0821, M08211, M08212, M08219, M0822, M08221, M08222, M08229, M0823, M08231, M08232, M08239, M0824, M08241, M08242, M08249, M0825, M08251, M08252, M08259, M0826, M08261, M08262, M08269, M0827, M08271, M08272, M08279, M0828, M0829, M082A, M083, M084, M0840, M0841, M08411, M08412, M08419, M0842, M08421, M08422, M08429, M0843, M08431, M08432, M08439, M0844, M08441, M08442, M08449, M0845, M08451, M08452, M08459, M0846, M08461, M08462, M08469, M0847, M08471, M08472, M08479, M0848, M084A, M088, M0880, M0881, M08811, M08812, M08819, M0882, M08821, M08822, M08829, M0883, M08831, M08832, M08839, M0884, M08841, M08842, M08849, M0885, M08851, M08852, M08859, M0886, M08861, M08862, M08869, M0887, M08871, M08872, M08879, M0888, M0889, M089, M0890, M0891, M08911, M08912, M08919, M0892, M08921, M08922, M08929, M0893, M08931, M08932, M08939, M0894, M08941, M08942, M08949, M0895, M08951, M08952, M08959, M0896, M08961, M08962, M08969, M0897, M08971, M08972, M08979, M0898, M0899, M089A, M1A, M1A0, M1A00, M1A00X0, M1A00X1, M1A01, M1A011, M1A0110, M1A0111, M1A012, M1A0120, M1A0121, M1A019, M1A0190, M1A0191, M1A02, M1A021, M1A0210, M1A0211, M1A022, M1A0220, M1A0221, M1A029, M1A0290, M1A0291, M1A03, M1A031, M1A0310, M1A0311, M1A032, M1A0320, M1A0321, M1A039, M1A0390, M1A0391, M1A04, M1A041, M1A0410, M1A0411, M1A042, M1A0420, M1A0421, M1A049, M1A0490, M1A0491, M1A05, M1A051, M1A0510, M1A0511, M1A052, M1A0520, M1A0521, M1A059, M1A0590, M1A0591, M1A06, M1A061, M1A0610, M1A0611, M1A062, M1A0620, M1A0621, M1A069, M1A0690, M1A0691, M1A07, M1A071, M1A0710, M1A0711, M1A072, M1A0720, M1A0721, M1A079, M1A0790, M1A0791, M1A08, M1A08X0, M1A08X1, M1A09, M1A09X0, M1A09X1, M1A1, M1A10, M1A10X0, M1A10X1, M1A11, M1A111, M1A1110, M1A1111, M1A112, M1A1120, M1A1121, M1A119, M1A1190, M1A1191, M1A12, M1A121, M1A1210, M1A1211, M1A122, M1A1220, M1A1221, M1A129, M1A1290, M1A1291, M1A13, M1A131, M1A1310, M1A1311, M1A132, M1A1320, M1A1321, M1A139, M1A1390, M1A1391, M1A14, M1A141, M1A1410, M1A1411, M1A142, M1A1420, M1A1421, M1A149, M1A1490, M1A1491, M1A15, M1A151, M1A1510, M1A1511, M1A152, M1A1520, M1A1521, M1A159, M1A1590, M1A1591, M1A16, M1A161, M1A1610, M1A1611, M1A162, M1A1620, M1A1621, M1A169, M1A1690, M1A1691, M1A17, M1A171, M1A1710, M1A1711, M1A172, M1A1720, M1A1721, M1A179, M1A1790, M1A1791, M1A18, M1A18X0, M1A18X1, M1A19, M1A19X0, M1A19X1, M1A2, M1A20, M1A20X0, M1A20X1, M1A21, M1A211, M1A2110, M1A2111, M1A212, M1A2120, M1A2121, M1A219, M1A2190, M1A2191, M1A22, M1A221, M1A2210, M1A2211, M1A222, M1A2220, M1A2221, M1A229, M1A2290, M1A2291, M1A23, M1A231, M1A2310, M1A2311, M1A232, M1A2320, M1A2321, M1A239, M1A2390, M1A2391, M1A24, M1A241, M1A2410, M1A2411, M1A242, M1A2420, M1A2421, M1A249, M1A2490, M1A2491, M1A25, M1A251, M1A2510, M1A2511, M1A252, M1A2520, M1A2521, M1A259, M1A2590, M1A2591, M1A26, M1A261, M1A2610, M1A2611, M1A262, M1A2620, M1A2621, M1A269, M1A2690, M1A2691, M1A27, M1A271, M1A2710, M1A2711, M1A272, M1A2720, M1A2721, M1A279, M1A2790, M1A2791, M1A28, M1A28X0, M1A28X1, M1A29, M1A29X0, M1A29X1, M1A3, M1A30, M1A30X0, M1A30X1, M1A31, M1A311, M1A3110, M1A3111, M1A312, M1A3120, M1A3121, M1A319, M1A3190, M1A3191, M1A32, M1A321, M1A3210, M1A3211, M1A322, M1A3220, M1A3221, M1A329, M1A3290, M1A3291, M1A33, M1A331, M1A3310, M1A3311, M1A332, M1A3320, M1A3321, M1A339, M1A3390, M1A3391, M1A34, M1A341, M1A3410, M1A3411, M1A342, M1A3420, M1A3421, M1A349, M1A3490, M1A3491, M1A35, M1A351, M1A3510, M1A3511, M1A352, M1A3520, M1A3521, M1A359, M1A3590, M1A3591, M1A36, M1A361, M1A3610, M1A3611, M1A362, M1A3620, M1A3621, M1A369, M1A3690, M1A3691, M1A37, M1A371, M1A3710, M1A3711, M1A372, M1A3720, M1A3721, M1A379, M1A3790, M1A3791, M1A38, M1A38X0, M1A38X1, M1A39, M1A39X0, M1A39X1, M1A4, M1A40, M1A40X0, M1A40X1, M1A41, M1A411, M1A4110, M1A4111, M1A412, M1A4120, M1A4121, M1A419, M1A4190, M1A4191, M1A42, M1A421, M1A4210, M1A4211, M1A422, M1A4220, M1A4221, M1A429, M1A4290, M1A4291, M1A43, M1A431, M1A4310, M1A4311, M1A432, M1A4320, M1A4321, M1A439, M1A4390, M1A4391, M1A44, M1A441, M1A4410, M1A4411, M1A442, M1A4420, M1A4421, M1A449, M1A4490, M1A4491, M1A45, M1A451, M1A4510, M1A4511, M1A452, M1A4520, M1A4521, M1A459, M1A4590, M1A4591, M1A46, M1A461, M1A4610, M1A4611, M1A462, M1A4620, M1A4621, M1A469, M1A4690, M1A4691, M1A47, M1A471, M1A4710, M1A4711, M1A472, M1A4720, M1A4721, M1A479, M1A4790, M1A4791, M1A48, M1A48X0, M1A48X1, M1A49, M1A49X0, M1A49X1, M1A9, M1A9XX0, M1A9XX1, M10, M100, M1000, M1001, M10011, M10012, M10019, M1002, M10021, M10022, M10029, M1003, M10031, M10032, M10039, M1004, M10041, M10042, M10049, M1005, M10051, M10052, M10059, M1006, M10061, M10062, M10069, M1007, M10071, M10072, M10079, M1008, M1009, M101, M1010, M1011, M10111, M10112, M10119, M1012, M10121, M10122, M10129, M1013, M10131, M10132, M10139, M1014, M10141, M10142, M10149, M1015, M10151, M10152, M10159, M1016, M10161, M10162, M10169, M1017, M10171, M10172, M10179, M1018, M1019, M102, M1020, M1021, M10211, M10212, M10219, M1022, M10221, M10222, M10229, M1023, M10231, M10232, M10239, M1024, M10241, M10242, M10249, M1025, M10251, M10252, M10259, M1026, M10261, M10262, M10269, M1027, M10271, M10272, M10279, M1028, M1029, M103, M1030, M1031, M10311, M10312, M10319, M1032, M10321, M10322, M10329, M1033, M10331, M10332, M10339, M1034, M10341, M10342, M10349, M1035, M10351, M10352, M10359, M1036, M10361, M10362, M10369, M1037, M10371, M10372, M10379, M1038, M1039, M104, M1040, M1041, M10411, M10412, M10419, M1042, M10421, M10422, M10429, M1043, M10431, M10432, M10439, M1044, M10441, M10442, M10449, M1045, M10451, M10452, M10459, M1046, M10461, M10462, M10469, M1047, M10471, M10472, M10479, M1048, M1049, M109, M11, M110, M1100, M1101, M11011, M11012, M11019, M1102, M11021, M11022, M11029, M1103, M11031, M11032, M11039, M1104, M11041, M11042, M11049, M1105, M11051, M11052, M11059, M1106, M11061, M11062, M11069, M1107, M11071, M11072, M11079, M1108, M1109, M111, M1110, M1111, M11111, M11112, M11119, M1112, M11121, M11122, M11129, M1113, M11131, M11132, M11139, M1114, M11141, M11142, M11149, M1115, M11151, M11152, M11159, M1116, M11161, M11162, M11169, M1117, M11171, M11172, M11179, M1118, M1119, M112, M1120, M1121, M11211, M11212, M11219, M1122, M11221, M11222, M11229, M1123, M11231, M11232, M11239, M1124, M11241, M11242, M11249, M1125, M11251, M11252, M11259, M1126, M11261, M11262, M11269, M1127, M11271, M11272, M11279, M1128, M1129, M118, M1180, M1181, M11811, M11812, M11819, M1182, M11821, M11822, M11829, M1183, M11831, M11832, M11839, M1184, M11841, M11842, M11849, M1185, M11851, M11852, M11859, M1186, M11861, M11862, M11869, M1187, M11871, M11872, M11879, M1188, M1189, M119, M12, M120, M1200, M1201, M12011, M12012, M12019, M1202, M12021, M12022, M12029, M1203, M12031, M12032, M12039, M1204, M12041, M12042, M12049, M1205, M12051, M12052, M12059, M1206, M12061, M12062, M12069, M1207, M12071, M12072, M12079, M1208, M1209, M121, M1210, M1211, M12111, M12112, M12119, M1212, M12121, M12122, M12129, M1213, M12131, M12132, M12139, M1214, M12141, M12142, M12149, M1215, M12151, M12152, M12159, M1216, M12161, M12162, M12169, M1217, M12171, M12172, M12179, M1218, M1219, M122, M1220, M1221, M12211, M12212, M12219, M1222, M12221, M12222, M12229, M1223, M12231, M12232, M12239, M1224, M12241, M12242, M12249, M1225, M12251, M12252, M12259, M1226, M12261, M12262, M12269, M1227, M12271, M12272, M12279, M1228, M1229, M123, M1230, M1231, M12311, M12312, M12319, M1232, M12321, M12322, M12329, M1233, M12331, M12332, M12339, M1234, M12341, M12342, M12349, M1235, M12351, M12352, M12359, M1236, M12361, M12362, M12369, M1237, M12371, M12372, M12379, M1238, M1239, M124, M1240, M1241, M12411, M12412, M12419, M1242, M12421, M12422, M12429, M1243, M12431, M12432, M12439, M1244, M12441, M12442, M12449, M1245, M12451, M12452, M12459, M1246, M12461, M12462, M12469, M1247, M12471, M12472, M12479, M1248, M1249, M125, M1250, M1251, M12511, M12512, M12519, M1252, M12521, M12522, M12529, M1253, M12531, M12532, M12539, M1254, M12541, M12542, M12549, M1255, M12551, M12552, M12559, M1256, M12561, M12562, M12569, M1257, M12571, M12572, M12579, M1258, M1259, M128, M1280, M1281, M12811, M12812, M12819, M1282, M12821, M12822, M12829, M1283, M12831, M12832, M12839, M1284, M12841, M12842, M12849, M1285, M12851, M12852, M12859, M1286, M12861, M12862, M12869, M1287, M12871, M12872, M12879, M1288, M1289, M129, M13, M130, M131, M1310, M1311, M13111, M13112, M13119, M1312, M13121, M13122, M13129, M1313, M13131, M13132, M13139, M1314, M13141, M13142, M13149, M1315, M13151, M13152, M13159, M1316, M13161, M13162, M13169, M1317, M13171, M13172, M13179, M138, M1380, M1381, M13811, M13812, M13819, M1382, M13821, M13822, M13829, M1383, M13831, M13832, M13839, M1384, M13841, M13842, M13849, M1385, M13851, M13852, M13859, M1386, M13861, M13862, M13869, M1387, M13871, M13872, M13879, M1388, M1389, M14, M146, M1460, M1461, M14611, M14612, M14619, M1462, M14621, M14622, M14629, M1463, M14631, M14632, M14639, M1464, M14641, M14642, M14649, M1465, M14651, M14652, M14659, M1466, M14661, M14662, M14669, M1467, M14671, M14672, M14679, M1468, M1469, M148, M1480, M1481, M14811, M14812, M14819, M1482, M14821, M14822, M14829, M1483, M14831, M14832, M14839, M1484, M14841, M14842, M14849, M1485, M14851, M14852, M14859, M1486, M14861, M14862, M14869, M1487, M14871, M14872, M14879, M1488, M1489, M15, M150, M151, M152, M153, M154, M158, M159, M16, M160, M161, M1610, M1611, M1612, M162, M163, M1630, M1631, M1632, M164, M165, M1650, M1651, M1652, M166, M167, M169, M17, M170, M171, M1710, M1711, M1712, M172, M173, M1730, M1731, M1732, M174, M175, M179, M18, M180, M181, M1810, M1811, M1812, M182, M183, M1830, M1831, M1832, M184, M185, M1850, M1851, M1852, M189, M19, M190, M1901, M19011, M19012, M19019, M1902, M19021, M19022, M19029, M1903, M19031, M19032, M19039, M1904, M19041, M19042, M19049, M1907, M19071, M19072, M19079, M1909, M191, M1911, M19111, M19112, M19119, M1912, M19121, M19122, M19129, M1913, M19131, M19132, M19139, M1914, M19141, M19142, M19149, M1917, M19171, M19172, M19179, M1919, M192, M1921, M19211, M19212, M19219, M1922, M19221, M19222, M19229, M1923, M19231, M19232, M19239, M1924, M19241, M19242, M19249, M1927, M19271, M19272, M19279, M1929, M199, M1990, M1991, M1992, M1993, L405, L4050, L4051, L4052, L4053, L4054, L4059, M80, M800, M8000, M8000XA, M8000XD, M8000XG, M8000XK, M8000XP, M8000XS, M8001, M80011, M80011A, M80011D, M80011G, M80011K, M80011P, M80011S, M80012, M80012A, M80012D, M80012G, M80012K, M80012P, M80012S, M80019, M80019A, M80019D, M80019G, M80019K, M80019P, M80019S, M8002, M80021, M80021A, M80021D, M80021G, M80021K, M80021P, M80021S, M80022, M80022A, M80022D, M80022G, M80022K, M80022P, M80022S, M80029, M80029A, M80029D, M80029G, M80029K, M80029P, M80029S, M8003, M80031, M80031A, M80031D, M80031G, M80031K, M80031P, M80031S, M80032, M80032A, M80032D, M80032G, M80032K, M80032P, M80032S, M80039, M80039A, M80039D, M80039G, M80039K, M80039P, M80039S, M8004, M80041, M80041A, M80041D, M80041G, M80041K, M80041P, M80041S, M80042, M80042A, M80042D, M80042G, M80042K, M80042P, M80042S, M80049, M80049A, M80049D, M80049G, M80049K, M80049P, M80049S, M8005, M80051, M80051A, M80051D, M80051G, M80051K, M80051P, M80051S, M80052, M80052A, M80052D, M80052G, M80052K, M80052P, M80052S, M80059, M80059A, M80059D, M80059G, M80059K, M80059P, M80059S, M8006, M80061, M80061A, M80061D, M80061G, M80061K, M80061P, M80061S, M80062, M80062A, M80062D, M80062G, M80062K, M80062P, M80062S, M80069, M80069A, M80069D, M80069G, M80069K, M80069P, M80069S, M8007, M80071, M80071A, M80071D, M80071G, M80071K, M80071P, M80071S, M80072, M80072A, M80072D, M80072G, M80072K, M80072P, M80072S, M80079, M80079A, M80079D, M80079G, M80079K, M80079P, M80079S, M8008, M8008XA, M8008XD, M8008XG, M8008XK, M8008XP, M8008XS, M800A, M800AXA, M800AXD, M800AXG, M800AXK, M800AXP, M800AXS, M808, M8080, M8080XA, M8080XD, M8080XG, M8080XK, M8080XP, M8080XS, M8081, M80811, M80811A, M80811D, M80811G, M80811K, M80811P, M80811S, M80812, M80812A, M80812D, M80812G, M80812K, M80812P, M80812S, M80819, M80819A, M80819D, M80819G, M80819K, M80819P, M80819S, M8082, M80821, M80821A, M80821D, M80821G, M80821K, M80821P, M80821S, M80822, M80822A, M80822D, M80822G, M80822K, M80822P, M80822S, M80829, M80829A, M80829D, M80829G, M80829K, M80829P, M80829S, M8083, M80831, M80831A, M80831D, M80831G, M80831K, M80831P, M80831S, M80832, M80832A, M80832D, M80832G, M80832K, M80832P, M80832S, M80839, M80839A, M80839D, M80839G, M80839K, M80839P, M80839S, M8084, M80841, M80841A, M80841D, M80841G, M80841K, M80841P, M80841S, M80842, M80842A, M80842D, M80842G, M80842K, M80842P, M80842S, M80849, M80849A, M80849D, M80849G, M80849K, M80849P, M80849S, M8085, M80851, M80851A, M80851D, M80851G, M80851K, M80851P, M80851S, M80852, M80852A, M80852D, M80852G, M80852K, M80852P, M80852S, M80859, M80859A, M80859D, M80859G, M80859K, M80859P, M80859S, M8086, M80861, M80861A, M80861D, M80861G, M80861K, M80861P, M80861S, M80862, M80862A, M80862D, M80862G, M80862K, M80862P, M80862S, M80869, M80869A, M80869D, M80869G, M80869K, M80869P, M80869S, M8087, M80871, M80871A, M80871D, M80871G, M80871K, M80871P, M80871S, M80872, M80872A, M80872D, M80872G, M80872K, M80872P, M80872S, M80879, M80879A, M80879D, M80879G, M80879K, M80879P, M80879S, M8088, M8088XA, M8088XD, M8088XG, M8088XK, M8088XP, M8088XS, M808A, M808AXA, M808AXD, M808AXG, M808AXK, M808AXP, M808AXS, M81, M810, M816, M818, M6284, M797 |
| Neurologic conditions | ICD-10-CM Dx | G1221, G40A1, G40A11, G40A19, G40B, G40B0, G40B01, G40B09, G40B1, G40B11, G40B19, G10, G91, G910, G911, G912, G913, G918, G919, G35, G71, G710, G7100, G7101, G7102, G7109, G711, G7111, G7112, G7113, G7114, G7119, G712, G7120, G7121, G7122, G71220, G71228, G7129, G713, G718, G719, G72, G720, G721, G722, G723, G724, G7241, G7249, G728, G7281, G7289, G729, G73, G731, G733, G737, G21, G210, G211, G2111, G2119, G212, G213, G214, G218, G219, G20 |
| Obesity (body mass index > 30) | ICD-10-CM Dx | E6601, E6609, E661, E662, E668, E669, Z6830, Z6831, Z6832, Z6833, Z6834, Z6835, Z6836, Z6837, Z6838, Z6839, Z6841, Z6842, Z6843, Z6844, Z6845 |
| Other immunocompromised condition | ICD-10-CM Dx | G113, D80, D800, D803, D804, D805, D806, D807, D81, D810, D811, D812, D815, D816, D817, D8189, D819, D82, D820, D821, D824, D83, D830, D831, D832, D838, D839, D8481, D822, D823, D828, D829, D84821, D849 |
| Physical inactivity | ICD-10-CM Dx | Z723 |
| Pregnancy (only in individuals 18+) | ICD-10-CM Dx | O00, O000, O0000, O0001, O001, O0010, O00101, O00102, O00109, O0011, O00111, O00112, O00119, O002, O0020, O00201, O00202, O00209, O0021, O00211, O00212, O00219, O008, O0080, O0081, O009, O0090, O0091, O02, O020, O021, O0281, O0289, O029, O03, O030, O031, O032, O033, O0330, O0331, O0332, O0333, O0334, O0335, O0336, O0337, O0338, O0339, O034, O035, O036, O037, O038, O0380, O0381, O0382, O0383, O0384, O0385, O0386, O0387, O0388, O0389, O039, O04, O045, O046, O047, O048, O0480, O0481, O0482, O0483, O0484, O0485, O0486, O0487, O0488, O0489, O0900, O0901, O0902, O0903, O0910, O0911, O0912, O0913, O09211, O09212, O09213, O09219, O09291, O09292, O09293, O09299, O0930, O0931, O0932, O0933, O0940, O0941, O0942, O0943, O09511, O09512, O09513, O09519, O09521, O09522, O09523, O09529, O09611, O09612, O09613, O09619, O09621, O09622, O09623, O09629, O0970, O0971, O0972, O0973, O09811, O09812, O09813, O09819, O09821, O09822, O09823, O09829, O09891, O09892, O09893, O09899, O0990, O0991, O0992, O0993, O09A0, O09A1, O09A2, O09A3, O10011, O10012, O10013, O10019, O1002, O10111, O10112, O10113, O10119, O1012, O10211, O10212, O10213, O10219, O1022, O10311, O10312, O10313, O10319, O1032, O10411, O10412, O10413, O10419, O1042, O10911, O10912, O10913, O10919, O1092, O111, O112, O113, O114, O119, O1200, O1201, O1202, O1203, O1204, O1210, O1211, O1212, O1213, O1214, O1220, O1221, O1222, O1223, O1224, O131, O132, O133, O134, O139, O1400, O1402, O1403, O1404, O1410, O1412, O1413, O1414, O1420, O1422, O1423, O1424, O1490, O1492, O1493, O1494, O1500, O1502, O1503, O161, O162, O163, O164, O169, O200, O208, O209, O210, O211, O212, O218, O219, O2200, O2201, O2202, O2203, O2210, O2211, O2212, O2213, O2220, O2221, O2222, O2223, O2230, O2231, O2232, O2233, O2240, O2241, O2242, O2243, O2250, O2251, O2252, O2253, O228X1, O228X2, O228X3, O228X9, O2290, O2291, O2292, O2293, O2300, O2301, O2302, O2303, O2310, O2311, O2312, O2313, O2320, O2321, O2322, O2323, O2330, O2331, O2332, O2333, O2340, O2341, O2342, O2343, O23511, O23512, O23513, O23519, O23521, O23522, O23523, O23529, O23591, O23592, O23593, O23599, O2390, O2391, O2392, O2393, O24011, O24012, O24013, O24019, O2402, O24111, O24112, O24113, O24119, O2412, O24311, O24312, O24313, O24319, O2432, O24410, O24414, O24415, O24419, O24420, O24424, O24425, O24429, O24811, O24812, O24813, O24819, O2482, O24911, O24912, O24913, O24919, O2492, O2510, O2511, O2512, O2513, O252, O2600, O2601, O2602, O2603, O2610, O2611, O2612, O2613, O2620, O2621, O2622, O2623, O2630, O2631, O2632, O2633, O2640, O2641, O2642, O2643, O2650, O2651, O2652, O2653, O26611, O26612, O26613, O26619, O2662, O26711, O26712, O26713, O26719, O2672, O26811, O26812, O26813, O26819, O26821, O26822, O26823, O26829, O26831, O26832, O26833, O26839, O26841, O26842, O26843, O26849, O26851, O26852, O26853, O26859, O2686, O26872, O26873, O26879, O26891, O26892, O26893, O26899, O2690, O2691, O2692, O2693, O280, O281, O282, O283, O284, O285, O288, O289, O29011, O29012, O29013, O29019, O29021, O29022, O29023, O29029, O29091, O29092, O29093, O29099, O29111, O29112, O29113, O29119, O29121, O29122, O29123, O29129, O29191, O29192, O29193, O29199, O29211, O29212, O29213, O29219, O29291, O29292, O29293, O29299, O293X1, O293X2, O293X3, O293X9, O2940, O2941, O2942, O2943, O295X1, O295X2, O295X3, O295X9, O2960, O2961, O2962, O2963, O298X1, O298X2, O298X3, O298X9, O2990, O2991, O2992, O2993, O30001, O30002, O30003, O30009, O30011, O30012, O30013, O30019, O30021, O30022, O30023, O30029, O30031, O30032, O30033, O30039, O30041, O30042, O30043, O30049, O30091, O30092, O30093, O30099, O30101, O30102, O30103, O30109, O30111, O30112, O30113, O30119, O30121, O30122, O30123, O30129, O30191, O30192, O30193, O30199, O30201, O30202, O30203, O30209, O30211, O30212, O30213, O30219, O30221, O30222, O30223, O30229, O30291, O30292, O30293, O30299, O30801, O30802, O30803, O30809, O30811, O30812, O30813, O30819, O30821, O30822, O30823, O30829, O30891, O30892, O30893, O30899, O3090, O3091, O3092, O3093, O3100X0, O3100X1, O3100X2, O3100X3, O3100X4, O3100X5, O3100X9, O3101X0, O3101X1, O3101X2, O3101X3, O3101X4, O3101X5, O3101X9, O3102X0, O3102X1, O3102X2, O3102X3, O3102X4, O3102X5, O3102X9, O3103X0, O3103X1, O3103X2, O3103X3, O3103X4, O3103X5, O3103X9, O3110X0, O3110X1, O3110X2, O3110X3, O3110X4, O3110X5, O3110X9, O3111X0, O3111X1, O3111X2, O3111X3, O3111X4, O3111X5, O3111X9, O3112X0, O3112X1, O3112X2, O3112X3, O3112X4, O3112X5, O3112X9, O3113X0, O3113X1, O3113X2, O3113X3, O3113X4, O3113X5, O3113X9, O3120X0, O3120X1, O3120X2, O3120X3, O3120X4, O3120X5, O3120X9, O3121X0, O3121X1, O3121X2, O3121X3, O3121X4, O3121X5, O3121X9, O3122X0, O3122X1, O3122X2, O3122X3, O3122X4, O3122X5, O3122X9, O3123X0, O3123X1, O3123X2, O3123X3, O3123X4, O3123X5, O3123X9, O3130X0, O3130X1, O3130X2, O3130X3, O3130X4, O3130X5, O3130X9, O3131X0, O3131X1, O3131X2, O3131X3, O3131X4, O3131X5, O3131X9, O3132X0, O3132X1, O3132X2, O3132X3, O3132X4, O3132X5, O3132X9, O3133X0, O3133X1, O3133X2, O3133X3, O3133X4, O3133X5, O3133X9, O318X10, O318X11, O318X12, O318X13, O318X14, O318X15, O318X19, O318X20, O318X21, O318X22, O318X23, O318X24, O318X25, O318X29, O318X30, O318X31, O318X32, O318X33, O318X34, O318X35, O318X39, O318X90, O318X91, O318X92, O318X93, O318X94, O318X95, O318X99, O320XX0, O320XX1, O320XX2, O320XX3, O320XX4, O320XX5, O320XX9, O321XX0, O321XX1, O321XX2, O321XX3, O321XX4, O321XX5, O321XX9, O322XX0, O322XX1, O322XX2, O322XX3, O322XX4, O322XX5, O322XX9, O323XX0, O323XX1, O323XX2, O323XX3, O323XX4, O323XX5, O323XX9, O324XX0, O324XX1, O324XX2, O324XX3, O324XX4, O324XX5, O324XX9, O326XX0, O326XX1, O326XX2, O326XX3, O326XX4, O326XX5, O326XX9, O328XX0, O328XX1, O328XX2, O328XX3, O328XX4, O328XX5, O328XX9, O329XX0, O329XX1, O329XX2, O329XX3, O329XX4, O329XX5, O329XX9, O330, O331, O332, O333XX0, O333XX1, O333XX2, O333XX3, O333XX4, O333XX5, O333XX9, O334XX0, O334XX1, O334XX2, O334XX3, O334XX4, O334XX5, O334XX9, O335XX0, O335XX1, O335XX2, O335XX3, O335XX4, O335XX5, O335XX9, O336XX0, O336XX1, O336XX2, O336XX3, O336XX4, O336XX5, O336XX9, O337XX0, O337XX1, O337XX2, O337XX3, O337XX4, O337XX5, O337XX9, O338, O339, O3400, O3401, O3402, O3403, O3410, O3411, O3412, O3413, O34211, O34212, O34219, O3429, O3430, O3431, O3432, O3433, O3440, O3441, O3442, O3443, O34511, O34512, O34513, O34519, O34521, O34522, O34523, O34529, O34531, O34532, O34533, O34539, O34591, O34592, O34593, O34599, O3460, O3461, O3462, O3463, O3470, O3471, O3472, O3473, O3480, O3481, O3482, O3483, O3490, O3491, O3492, O3493, O350XX0, O350XX1, O350XX2, O350XX3, O350XX4, O350XX5, O350XX9, O351XX0, O351XX1, O351XX2, O351XX3, O351XX4, O351XX5, O351XX9, O352XX0, O352XX1, O352XX2, O352XX3, O352XX4, O352XX5, O352XX9, O353XX0, O353XX1, O353XX2, O353XX3, O353XX4, O353XX5, O353XX9, O354XX0, O354XX1, O354XX2, O354XX3, O354XX4, O354XX5, O354XX9, O355XX0, O355XX1, O355XX2, O355XX3, O355XX4, O355XX5, O355XX9, O356XX0, O356XX1, O356XX2, O356XX3, O356XX4, O356XX5, O356XX9, O357XX0, O357XX1, O357XX2, O357XX3, O357XX4, O357XX5, O357XX9, O358XX0, O358XX1, O358XX2, O358XX3, O358XX4, O358XX5, O358XX9, O359XX0, O359XX1, O359XX2, O359XX3, O359XX4, O359XX5, O359XX9, O360110, O360111, O360112, O360113, O360114, O360115, O360119, O360120, O360121, O360122, O360123, O360124, O360125, O360129, O360130, O360131, O360132, O360133, O360134, O360135, O360139, O360190, O360191, O360192, O360193, O360194, O360195, O360199, O360910, O360911, O360912, O360913, O360914, O360915, O360919, O360920, O360921, O360922, O360923, O360924, O360925, O360929, O360930, O360931, O360932, O360933, O360934, O360935, O360939, O360990, O360991, O360992, O360993, O360994, O360995, O360999, O361110, O361111, O361112, O361113, O361114, O361115, O361119, O361120, O361121, O361122, O361123, O361124, O361125, O361129, O361130, O361131, O361132, O361133, O361134, O361135, O361139, O361190, O361191, O361192, O361193, O361194, O361195, O361199, O361910, O361911, O361912, O361913, O361914, O361915, O361919, O361920, O361921, O361922, O361923, O361924, O361925, O361929, O361930, O361931, O361932, O361933, O361934, O361935, O361939, O361990, O361991, O361992, O361993, O361994, O361995, O361999, O3620X0, O3620X1, O3620X2, O3620X3, O3620X4, O3620X5, O3620X9, O3621X0, O3621X1, O3621X2, O3621X3, O3621X4, O3621X5, O3621X9, O3622X0, O3622X1, O3622X2, O3622X3, O3622X4, O3622X5, O3622X9, O3623X0, O3623X1, O3623X2, O3623X3, O3623X4, O3623X5, O3623X9, O364, O364XX0, O364XX1, O364XX2, O364XX5, O364XX9, O365110, O365111, O365112, O365113, O365114, O365115, O365119, O365120, O365121, O365122, O365123, O365124, O365125, O365129, O365130, O365131, O365132, O365133, O365134, O365135, O365139, O365190, O365191, O365192, O365193, O365194, O365195, O365199, O365910, O365911, O365912, O365913, O365914, O365915, O365919, O365920, O365921, O365922, O365923, O365924, O365925, O365929, O365930, O365931, O365932, O365933, O365934, O365935, O365939, O365990, O365991, O365992, O365993, O365994, O365995, O365999, O3660X0, O3660X1, O3660X2, O3660X3, O3660X4, O3660X5, O3660X9, O3661X0, O3661X1, O3661X2, O3661X3, O3661X4, O3661X5, O3661X9, O3662X0, O3662X1, O3662X2, O3662X3, O3662X4, O3662X5, O3662X9, O3663X0, O3663X1, O3663X2, O3663X3, O3663X4, O3663X5, O3663X9, O3670X0, O3670X1, O3670X2, O3670X3, O3670X4, O3670X5, O3670X9, O3671X0, O3671X1, O3671X2, O3671X3, O3671X4, O3671X5, O3671X9, O3672X0, O3672X1, O3672X2, O3672X3, O3672X4, O3672X5, O3672X9, O3673X0, O3673X1, O3673X2, O3673X3, O3673X4, O3673X5, O3673X9, O368120, O368121, O368122, O368123, O368124, O368125, O368129, O368130, O368131, O368132, O368133, O368134, O368135, O368139, O368190, O368191, O368192, O368193, O368194, O368195, O368199, O368210, O368211, O368212, O368213, O368214, O368215, O368219, O368220, O368221, O368222, O368223, O368224, O368225, O368229, O368230, O368231, O368232, O368233, O368234, O368235, O368239, O368290, O368291, O368292, O368293, O368294, O368295, O368299, O368910, O368911, O368912, O368913, O368914, O368915, O368919, O368920, O368921, O368922, O368923, O368924, O368925, O368929, O368930, O368931, O368932, O368933, O368934, O368935, O368939, O368990, O368991, O368992, O368993, O368994, O368995, O368999, O3690X0, O3690X1, O3690X2, O3690X3, O3690X4, O3690X5, O3690X9, O3691X0, O3691X1, O3691X2, O3691X3, O3691X4, O3691X5, O3691X9, O3692X0, O3692X1, O3692X2, O3692X3, O3692X4, O3692X5, O3692X9, O3693X0, O3693X1, O3693X2, O3693X3, O3693X4, O3693X5, O3693X9, O401XX0, O401XX1, O401XX2, O401XX3, O401XX4, O401XX5, O401XX9, O402XX0, O402XX1, O402XX2, O402XX3, O402XX4, O402XX5, O402XX9, O403XX0, O403XX1, O403XX2, O403XX3, O403XX4, O403XX5, O403XX9, O409XX0, O409XX1, O409XX2, O409XX3, O409XX4, O409XX5, O409XX9, O4100X0, O4100X1, O4100X2, O4100X3, O4100X4, O4100X5, O4100X9, O4101X0, O4101X1, O4101X2, O4101X3, O4101X4, O4101X5, O4101X9, O4102X0, O4102X1, O4102X2, O4102X3, O4102X4, O4102X5, O4102X9, O4103X0, O4103X1, O4103X2, O4103X3, O4103X4, O4103X5, O4103X9, O411010, O411011, O411012, O411013, O411014, O411015, O411019, O411020, O411021, O411022, O411023, O411024, O411025, O411029, O411030, O411031, O411032, O411033, O411034, O411035, O411039, O411090, O411091, O411092, O411093, O411094, O411095, O411099, O411210, O411211, O411212, O411213, O411214, O411215, O411219, O411220, O411221, O411222, O411223, O411224, O411225, O411229, O411230, O411231, O411232, O411233, O411234, O411235, O411239, O411290, O411291, O411292, O411293, O411294, O411295, O411299, O411410, O411411, O411412, O411413, O411414, O411415, O411419, O411420, O411421, O411422, O411423, O411424, O411425, O411429, O411430, O411431, O411432, O411433, O411434, O411435, O411439, O411490, O411491, O411492, O411493, O411494, O411495, O411499, O418X10, O418X11, O418X12, O418X13, O418X14, O418X15, O418X19, O418X20, O418X21, O418X22, O418X23, O418X24, O418X25, O418X29, O418X30, O418X31, O418X32, O418X33, O418X34, O418X35, O418X39, O418X90, O418X91, O418X92, O418X93, O418X94, O418X95, O418X99, O4190X0, O4190X1, O4190X2, O4190X3, O4190X4, O4190X5, O4190X9, O4191X0, O4191X1, O4191X2, O4191X3, O4191X4, O4191X5, O4191X9, O4192X0, O4192X1, O4192X2, O4192X3, O4192X4, O4192X5, O4192X9, O4193X0, O4193X1, O4193X2, O4193X3, O4193X4, O4193X5, O4193X9, O4200, O42011, O42012, O42013, O42019, O4202, O4210, O42111, O42112, O42113, O42119, O4212, O4290, O42911, O42912, O42913, O42919, O4292, O43011, O43012, O43013, O43019, O43021, O43022, O43023, O43029, O43101, O43102, O43103, O43109, O43111, O43112, O43113, O43119, O43121, O43122, O43123, O43129, O43191, O43192, O43193, O43199, O43211, O43212, O43213, O43219, O43221, O43222, O43223, O43229, O43231, O43232, O43233, O43239, O43811, O43812, O43813, O43819, O43891, O43892, O43893, O43899, O4390, O4391, O4392, O4393, O4400, O4401, O4402, O4403, O4410, O4411, O4412, O4413, O4420, O4421, O4422, O4423, O4430, O4431, O4432, O4433, O4440, O4441, O4442, O4443, O4450, O4451, O4452, O4453, O45001, O45002, O45003, O45009, O45011, O45012, O45013, O45019, O45021, O45022, O45023, O45029, O45091, O45092, O45093, O45099, O458X1, O458X2, O458X3, O458X9, O4590, O4591, O4592, O4593, O46001, O46002, O46003, O46009, O46011, O46012, O46013, O46019, O46021, O46022, O46023, O46029, O46091, O46092, O46093, O46099, O468X1, O468X2, O468X3, O468X9, O4690, O4691, O4692, O4693, O4700, O4702, O4703, O471, O479, O480, O481, O6000, O6002, O6003, O6012, O6012X0, O6012X1, O6012X2, O6012X3, O6012X4, O6012X5, O6012X9, O6013, O6013X0, O6013X1, O6013X2, O6013X3, O6013X4, O6013X5, O6013X9, O6014, O6014X0, O6014X1, O6014X2, O6014X3, O6014X4, O6014X5, O6014X9, O6022, O6022X0, O6022X1, O6022X2, O6022X3, O6022X4, O6022X5, O6022X9, O6023, O6023X0, O6023X1, O6023X2, O6023X3, O6023X4, O6023X5, O6023X9, O632, O665, O67, O670, O678, O679, O68, O690, O690XX0, O690XX1, O690XX2, O690XX3, O690XX4, O690XX5, O690XX9, O691, O691XX0, O691XX1, O691XX2, O691XX3, O691XX4, O691XX5, O691XX9, O692, O692XX0, O692XX1, O692XX2, O692XX3, O692XX4, O692XX5, O692XX9, O693, O693XX0, O693XX1, O693XX2, O693XX3, O693XX4, O693XX5, O693XX9, O694, O694XX0, O694XX1, O694XX2, O694XX3, O694XX4, O694XX5, O694XX9, O695, O695XX0, O695XX1, O695XX2, O695XX3, O695XX4, O695XX5, O695XX9, O6981, O6981X0, O6981X1, O6981X2, O6981X3, O6981X4, O6981X5, O6981X9, O6982, O6982X0, O6982X1, O6982X2, O6982X3, O6982X4, O6982X5, O6982X9, O6989, O6989X0, O6989X1, O6989X2, O6989X3, O6989X4, O6989X5, O6989X9, O699, O699XX0, O699XX1, O699XX2, O699XX3, O699XX4, O699XX5, O699XX9, O70, O700, O701, O702, O7020, O7021, O7022, O7023, O703, O704, O709, O7100, O7102, O7103, O74, O740, O741, O742, O743, O744, O745, O746, O747, O748, O749, O750, O751, O755, O758, O7581, O7582, O7589, O759, O76, O77, O770, O771, O778, O779, O80, O82, O88011, O88012, O88013, O88019, O8802, O88111, O88112, O88113, O88119, O8812, O88211, O88212, O88213, O88219, O8822, O88311, O88312, O88313, O88319, O8832, O88811, O88812, O88813, O88819, O8882, O903, O91011, O91012, O91013, O91019, O91111, O91112, O91113, O91119, O91211, O91212, O91213, O91219, O92011, O92012, O92013, O92019, O92111, O92112, O92113, O92119, O9220, O9229, O98011, O98012, O98013, O98019, O9802, O98111, O98112, O98113, O98119, O9812, O98211, O98212, O98213, O98219, O9822, O98311, O98312, O98313, O98319, O9832, O98411, O98412, O98413, O98419, O9842, O98511, O98512, O98513, O98519, O9852, O98611, O98612, O98613, O98619, O9862, O98711, O98712, O98713, O98719, O9872, O98811, O98812, O98813, O98819, O9882, O98911, O98912, O98913, O98919, O9892, O99011, O99012, O99013, O99019, O9902, O99111, O99112, O99113, O99119, O9912, O99210, O99211, O99212, O99213, O99214, O99280, O99281, O99282, O99283, O99284, O99310, O99311, O99312, O99313, O99314, O99320, O99321, O99322, O99323, O99324, O99330, O99331, O99332, O99333, O99334, O99340, O99341, O99342, O99343, O99344, O99350, O99351, O99352, O99353, O99354, O99411, O99412, O99413, O99419, O9942, O99511, O99512, O99513, O99519, O9952, O99611, O99612, O99613, O99619, O9962, O99711, O99712, O99713, O99719, O9972, O99810, O99814, O99820, O99824, O99830, O99834, O99840, O99841, O99842, O99843, O99844, O9989, O9A111, O9A112, O9A113, O9A119, O9A12, O9A211, O9A212, O9A213, O9A219, O9A22, O9A311, O9A312, O9A313, O9A319, O9A32, O9A411, O9A412, O9A413, O9A419, O9A42, O9A511, O9A512, O9A513, O9A519, O9A52, P030, P032, P033, P034, P035, P0700, P0701, P0702, P0703, P0710, P0714, P0715, P0716, P0717, P0718, P0720, P0721, P0722, P0723, P0724, P0725, P0726, P0730, P0731, P0732, P0733, P0734, P0735, P0736, P0737, P0738, P0739, P0821, P0822, Z3201, Z331, Z332, Z333, Z34, Z340, Z3400, Z3401, Z3402, Z3403, Z348, Z3480, Z3481, Z3482, Z3483, Z349, Z3490, Z3491, Z3492, Z3493, Z361, Z3682, Z370, Z371, Z372, Z373, Z374, Z375, Z3750, Z3751, Z3752, Z3753, Z3754, Z3759, Z376, Z3760, Z3761, Z3762, Z3763, Z3764, Z3769, Z377, Z379, Z38, Z380, Z3800, Z3801, Z381, Z382, Z383, Z3830, Z3831, Z384, Z385, Z386, Z3861, Z3862, Z3863, Z3864, Z3865, Z3866, Z3868, Z3869, Z387, Z388, Z640, O310, O3100, O3101, O3102, O3103, O364XX3, O364XX4 |
| Smoking (current and former; only in individuals 18+) | ICD-10-CM Dx | F17, F172, F1720, F17200, F17201, F17203, F17208, F17209, F1721, F17210, F17211, F17213, F17218, F17219, F1722, F17220, F17221, F17223, F17228, F17229, F1729, F17290, F17291, F17293, F17298, F17299, Z720, Z87891, O9933, O99330, O99331, O99332, O99333, O99334, O99335 |
| Solid organ transplant | CPT | 32851, 32852, 32853, 32854, 33935, 33945, 44135, 44136, 47135, 47136, 48554, 50360, 50365, 50370 |
|  | DRG | 1, 2, 5, 6, 7, 8, 19, 650, 651, 652 |
|  | HCPCS | S2053, S2054, S2060, S2152, S2065 |
|  | ICD-10-CM Dx | Z4821, Z4822, Z4823, Z4824, Z48280, Z48288, Z940, Z941, Z942, Z943, Z944, Z9482, Z9483, T8610, T8619, T8620, T86298, T8630, T8639, T8640, T8649, T86818, T86819, T86858, T86859, T86898, T86899 |
|  | ICD-10-PCS | 02YA0Z0, 02YA0Z1, 02YA0Z2, 07YM0Z0, 07YM0Z1, 07YM0Z2, 07YP0Z0, 07YP0Z1, 07YP0Z2, 0BYC0Z0, 0BYC0Z1, 0BYC0Z2, 0BYD0Z0, 0BYD0Z1, 0BYD0Z2, 0BYF0Z0, 0BYF0Z1, 0BYF0Z2, 0BYH0Z0, 0BYH0Z1, 0BYH0Z2, 0BYJ0Z0, 0BYJ0Z1, 0BYJ0Z2, 0BYK0Z0, 0BYK0Z1, 0BYK0Z2, 0BYL0Z0, 0BYL0Z1, 0BYL0Z2, 0BYM0Z0, 0BYM0Z1, 0BYM0Z2, 0DY50Z0, 0DY50Z1, 0DY50Z2, 0DY60Z0, 0DY60Z1, 0DY60Z2, 0DY80Z0, 0DY80Z1, 0DY80Z2, 0DYE0Z0, 0DYE0Z1, 0DYE0Z2, 0FY00Z0, 0FY00Z1, 0FY00Z2, 0FYG0Z0, 0FYG0Z1, 0FYG0Z2, 0TY00Z0, 0TY00Z1, 0TY00Z2, 0TY10Z0, 0TY10Z1, 0TY10Z2, BT2900Z, BT290ZZ, BT2910Z, BT291ZZ, BT29Y0Z, BT29YZZ, BT29ZZZ, BT39Y0Z, BT39YZZ, BT39ZZZ, BT49ZZZ |
| Stem cell transplant | CPT | 38240, 38241, 38242, 38243 |
|  | DRG | 014, 016, 017 |
|  | HCPCS | S2142, S2150 |
|  | ICD-10-CM Dx | T8600, T8609, Z48290, Z9481 |
|  | ICD-10-PCS | 30230AZ, 30230G1, 30230G2, 30230G3, 30230G4, 30230X1, 30230X2, 30230X3, 30230X4, 30230Y1, 30230Y2, 30230Y3, 30230Y4, 30233AZ, 30233G1, 30233G2, 30233G3, 30233G4, 30233X1, 30233X2, 30233X3, 30233X4, 30233Y1, 30233Y2, 30233Y3, 30233Y4, 30240AZ, 30240G1, 30240G2, 30240G3, 30240G4, 30240X1, 30240X2, 30240X3, 30240X4, 30240Y1, 30240Y2, 30240Y3, 30240Y4, 30243AZ, 30243G1, 30243G2, 30243G3, 30243G4, 30243X1, 30243X2, 30243X3, 30243X4, 30243Y1, 30243Y2, 30243Y3, 30243Y4, 30250G1, 30250X1, 30250Y1, 30253G1, 30253X1, 30253Y1, 30260G1, 30260X1, 30260Y1, 30263G1, 30263X1, 30263Y1 |
| Tuberculosis | ICD-10-CM Dx | A150, A154, A155, A156, A157, A158, A159 |
