## Supplementary File 2 for "Comparison of COVID-19 and Influenza-Related Outcomes in the United States during Fall-Winter 2022-2023"

Supplementary File 2. Comorbidity burden among patients hospitalized with COVID-19

|  | Age 0-5 | | | |
| --- | --- | --- | --- | --- |
|  | Hospitalized* patients | | All individuals in dataset | |
|  | N = | 706 | N = | 506,690 |
|  | N | % | N | % |
| No comorbidity | 182 | 25.8% | 247,846 | 48.9% |
| Cancer | 59 | 8.4% | 11,012 | 2.2% |
| Cerebrovascular disease | 15 | 2.1% | 1,204 | 0.2% |
| Chronic kidney disease | 11 | 1.6% | 422 | 0.1% |
| Chronic lung disease | 78 | 11.0% | 3,160 | 0.6% |
| Chronic liver disease | <10 | N/A | 58 | 0.0% |
| Cystic fibrosis | <10 | N/A | 413 | 0.1% |
| Diabetes | <10 | N/A | 710 | 0.1% |
| Disability | 196 | 27.8% | 77,142 | 15.2% |
| Heart Condition | 18 | 2.5% | 1,062 | 0.2% |
| HIV | <10 | N/A | 41 | 0.0% |
| Mental health disorders | <10 | N/A | 702 | 0.1% |
| Dementia | N/A | N/A | N/A | N/A |
| Obesity (BMI > 30) | 10 | 1.4% | 7,966 | 1.6% |
| Other Immunodeficiencies | 47 | 6.7% | 3,962 | 0.8% |
| Pregnancy | N/A | N/A | N/A | N/A |
| Physical inactivity | <10 | N/A | 183 | 0.0% |
| Smoking (current and former) | N/A | N/A | N/A | N/A |
| Solid organ transplant | <10 | N/A | 120 | 0.0% |
| Tuberculosis | <10 | N/A | 30 | 0.0% |
| Immunosuppressive medication | 349 | 49.4% | 162,315 | 32.0% |
| Asthma | 151 | 21.4% | 40,489 | 8.0% |
| Neurologic conditions | 23 | 3.3% | 1,166 | 0.2% |
| Musculoskeletal conditions | <10 | N/A | 377 | 0.1% |
| Hypertension | 37 | 5.2% | 1,445 | 0.3% |
| ADHD | <10 | N/A | 4,291 | 0.8% |
| Cerebral palsy | 29 | 4.1% | 1,480 | 0.3% |
| Congenital malformation | 296 | 41.9% | 68,519 | 13.5% |
| Down syndrome | 14 | 2.0% | 1,242 | 0.2% |
| Stem cell transplant | <10 | N/A | 67 | 0.0% |

*cell counts of 1-9 are reported as <10 to ensure data is deidentified. Zeros are reported as zeros.

ADHD, attention deficit hyperactivity disorder; BMI, body mass index; HIV, human immunodeficiency virus

|  | Age 6-17 | | | |
| --- | --- | --- | --- | --- |
|  | Hospitalized* patients | | All individuals in dataset | |
|  | N = | 1,529 | N = | 2,794,168 |
|  | N | % | N | % |
| No comorbidity | 280 | 18.3% | 1,426,772 | 51.1% |
| Cancer | 155 | 10.1% | 64,481 | 2.3% |
| Cerebrovascular disease | 48 | 3.1% | 3,350 | 0.1% |
| Chronic kidney disease | 43 | 2.8% | 2,972 | 0.1% |
| Chronic lung disease | 108 | 7.1% | 5,427 | 0.2% |
| Chronic liver disease | 10 | 0.7% | 1,136 | 0.0% |
| Cystic fibrosis | 14 | 0.9% | 1,214 | 0.0% |
| Diabetes | 69 | 4.5% | 20,212 | 0.7% |
| Disability | 411 | 26.9% | 237,885 | 8.5% |
| Heart Condition | 53 | 3.5% | 3,976 | 0.1% |
| HIV | <10 | N/A | 368 | 0.0% |
| Mental health disorders | 336 | 22.0% | 174,464 | 6.2% |
| Dementia | N/A | N/A | N/A | N/A |
| Obesity (BMI > 30) | 174 | 11.4% | 175,954 | 6.3% |
| Other Immunodeficiencies | 148 | 9.7% | 6,162 | 0.2% |
| Pregnancy | N/A | N/A | N/A | N/A |
| Physical inactivity | <10 | N/A | 1,871 | 0.1% |
| Smoking (current and former) | N/A | N/A | N/A | N/A |
| Solid organ transplant | 30 | 2.0% | 1,250 | 0.0% |
| Tuberculosis | 0 | 0.0% | 137 | 0.0% |
| Immunosuppressive medication | 736 | 48.1% | 729,922 | 26.1% |
| Asthma | 394 | 25.8% | 303,733 | 10.9% |
| Neurologic conditions | 56 | 3.7% | 5,672 | 0.2% |
| Musculoskeletal conditions | 26 | 1.7% | 7,961 | 0.3% |
| Hypertension | 118 | 7.7% | 17,596 | 0.6% |
| ADHD | 315 | 20.6% | 354,833 | 12.7% |
| Cerebral palsy | 143 | 9.4% | 10,353 | 0.4% |
| Congenital malformation | 350 | 22.9% | 130,976 | 4.7% |
| Down syndrome | 16 | 1.0% | 5,749 | 0.2% |
| Stem cell transplant | 12 | 0.8% | 555 | 0.0% |

*cell counts of 1-9 are reported as <10 to ensure data is deidentified. Zeros are reported as zeros.

ADHD, attention deficit hyperactivity disorder; BMI, body mass index; HIV, human immunodeficiency virus

|  | Age 18-64 | | | |
| --- | --- | --- | --- | --- |
|  | Hospitalized* patients | | All individuals in dataset | |
|  | N = | 49,189 | N = | 15,454,879 |
|  | N | % | N | % |
| No comorbidity | 2,779 | 5.6% | 4,450,082 | 28.8% |
| Cancer | 11,428 | 23.2% | 2,385,924 | 15.4% |
| Cerebrovascular disease | 5,063 | 10.3% | 292,406 | 1.9% |
| Chronic kidney disease | 7,805 | 15.9% | 347,900 | 2.3% |
| Chronic lung disease | 10,787 | 21.9% | 525,818 | 3.4% |
| Chronic liver disease | 2,306 | 4.7% | 121,708 | 0.8% |
| Cystic fibrosis | 89 | 0.2% | 3,910 | 0.0% |
| Diabetes | 15,147 | 30.8% | 1,762,550 | 11.4% |
| Disability | 1,131 | 2.3% | 153,525 | 1.0% |
| Heart Condition | 11,987 | 24.4% | 727,435 | 4.7% |
| HIV | 824 | 1.7% | 87,633 | 0.6% |
| Mental health disorders | 16,459 | 33.5% | 2,572,057 | 16.6% |
| Dementia | 1,605 | 3.3% | 52,931 | 0.3% |
| Obesity (BMI > 30) | 19,831 | 40.3% | 3,543,618 | 22.9% |
| Other Immunodeficiencies | 2,187 | 4.4% | 85,959 | 0.6% |
| Pregnancy | 9,901 | 20.1% | 655,765 | 4.2% |
| Physical inactivity | 139 | 0.3% | 12,625 | 0.1% |
| Smoking (current and former) | 17,657 | 35.9% | 1,999,052 | 12.9% |
| Solid organ transplant | 1,320 | 2.7% | 36,212 | 0.2% |
| Tuberculosis | 74 | 0.2% | 4,798 | 0.0% |
| Immunosuppressive medication | 24,109 | 49.0% | 5,372,488 | 34.8% |
| Asthma | 9,546 | 19.4% | 1,278,488 | 8.3% |
| Neurologic conditions | 1,782 | 3.6% | 117,760 | 0.8% |
| Musculoskeletal conditions | 12,847 | 26.1% | 1,885,491 | 12.2% |
| Hypertension | 24,891 | 50.6% | 3,713,659 | 24.0% |
| ADHD | 2,402 | 4.9% | 757,056 | 4.9% |
| Cerebral palsy | 379 | 0.8% | 27,729 | 0.2% |
| Congenital malformation | 3,338 | 6.8% | 392,380 | 2.5% |
| Down syndrome | 95 | 0.2% | 10,815 | 0.1% |
| Stem cell transplant | 157 | 0.3% | 4,474 | 0.0% |

*cell counts of 1-9 are reported as <10 to ensure data is deidentified. Zeros are reported as zeros.

ADHD, attention deficit hyperactivity disorder; BMI, body mass index; HIV, human immunodeficiency virus

|  | Age 65+ | | | |
| --- | --- | --- | --- | --- |
|  | Hospitalized* patients | | All individuals in dataset | |
|  | N = | 42,464 | N = | 4,770,459 |
|  | N | % | N | % |
| No comorbidity | 1,130 | 0.4% | 458,448 | 9.6% |
| Cancer | 15,770 | 37.1% | 1,516,148 | 31.8% |
| Cerebrovascular disease | 10,995 | 25.9% | 510,463 | 10.7% |
| Chronic kidney disease | 15,699 | 37.0% | 760,873 | 15.9% |
| Chronic lung disease | 16,731 | 39.4% | 704,346 | 14.8% |
| Chronic liver disease | 1,517 | 3.6% | 69,276 | 1.5% |
| Cystic fibrosis | 13 | 0.0% | 441 | 0.0% |
| Diabetes | 20,931 | 49.3% | 1,471,256 | 30.8% |
| Disability | 319 | 0.8% | 14,801 | 0.3% |
| Heart Condition | 22,935 | 54.0% | 1,185,761 | 24.9% |
| HIV | 261 | 0.6% | 16,222 | 0.3% |
| Mental health disorders | 12,325 | 29.0% | 757,323 | 15.9% |
| Dementia | 8,802 | 20.7% | 314,610 | 6.6% |
| Obesity (BMI > 30) | 14,063 | 33.1% | 1,214,499 | 25.5% |
| Other Immunodeficiencies | 1,873 | 4.4% | 74,559 | 1.6% |
| Pregnancy | <10 | N/A | 185 | 0.0% |
| Physical inactivity | 143 | 0.3% | 5,943 | 0.1% |
| Smoking (current and former) | 13,963 | 32.9% | 803,421 | 16.8% |
| Solid organ transplant | 776 | 1.8% | 17,103 | 0.4% |
| Tuberculosis | 45 | 0.1% | 2,303 | 0.0% |
| Immunosuppressive medication | 22,385 | 52.7% | 2,091,105 | 43.8% |
| Asthma | 5,961 | 14.0% | 375,417 | 7.9% |
| Neurologic conditions | 2,803 | 6.6% | 113,066 | 2.4% |
| Musculoskeletal conditions | 22,864 | 53.8% | 1,878,269 | 39.4% |
| Hypertension | 36,246 | 85.4% | 3,091,563 | 64.8% |
| ADHD | 209 | 0.5% | 23,642 | 0.5% |
| Cerebral palsy | 96 | 0.2% | 4,372 | 0.1% |
| Congenital malformation | 2,306 | 5.4% | 144,695 | 3.0% |
| Down syndrome | 13 | 0.0% | 357 | 0.0% |
| Stem cell transplant | 77 | 0.2% | 2,307 | 0.0% |

*cell counts of 1-9 are reported as <10 to ensure data is deidentified. Zeros are reported as zeros.

ADHD, attention deficit hyperactivity disorder; BMI, body mass index; HIV, human immunodeficiency virus
